# Investigating the Characteristics of Genes and Variants Associated with Self-Reported Hearing Difficulty in Older Adults in the UK Biobank

**DOI:** 10.1101/2022.01.28.22269991

**Authors:** Morag A. Lewis, Bradley A. Schulte, Judy R. Dubno, Karen P. Steel

## Abstract

**Background:** Age-related hearing loss is a common, heterogeneous disease with a strong genetic component. More than 100 loci have been reported to be involved in human hearing impairment to date, but most of the genes underlying human adult-onset hearing loss remain unknown. Most genetic studies have focussed on very rare variants (such as family studies and patient cohort screens) or very common variants (genome-wide association studies). However, the contribution of variants present in the human population at intermediate frequencies is hard to quantify using these methods, and as a result, the landscape of variation associated with adult-onset hearing loss remains largely unknown.

**Results:** Here we present a study based on exome sequencing and self-reported hearing difficulty in the UK Biobank, a large-scale biomedical database. We have carried out variant load analyses using different minor allele frequency and impact filters, and compared the resulting gene lists to a manually-curated list of nearly 700 genes known to be involved in hearing in humans and/or mice. An allele frequency cutoff of 0.1, combined with a high predicted variant impact, was found to be the most effective filter settings for our analysis. We also found that separating the participants by sex produced markedly different gene lists. The gene lists obtained were investigated using gene ontology annotation, functional prioritisation and expression analysis, and this identified good candidates for further study.

**Conclusions:** Our results suggest that relatively common as well as rare variants with a high predicted impact contribute to age-related hearing impairment, and that the genetic contributions to adult hearing difficulty may differ between the sexes. Our manually-curated list of deafness genes is a useful resource for candidate gene prioritisation in hearing loss.

## Background

Hearing impairment is one of the most common sensory deficits in the human population and has a strong genetic component. However, the auditory system is a complex system with many interacting parts, which offers many routes to loss of function. Accordingly, although over 150 genes have been identified as contributing to non-syndromic human hearing loss [1], the majority of genes involved in hearing remain unknown. Moreover, most of the genes identified so far are those where mutations result in early-onset, severe hearing loss. While age-related hearing loss (ARHL) is very common, it is also very heterogeneous, and the associated landscape of genetic variation remains unclear, both at the gene and at the variant level. Even when analysing rare variants in known deafness genes, a wide mutational spectrum can be observed, with a range of allele frequencies and predicted impacts which differ on a gene-by-gene basis [2].

As early as 1997 it was noted that single gene mutations can lead to early postnatal or adult-onset progressive hearing loss [3]. This remains the case 25 years later; 45 out of the 51 known human autosomal dominant deafness genes result in progressive hearing loss when mutated [1]. These mutations are rare, high-impact variants which have been identified through family studies and candidate gene screening of patient cohorts, for example [4-7]. However, most such variants are ultra-rare or even private [5], and while they fully explain the hearing loss seen in the affected individual or family, they cannot explain all the ARHL seen in the population. On the other end of the scale, looking at common variants, very large genome wide association studies (GWAS) have recently uncovered several new loci [8, 9], but because GWAS work by identifying markers linked to disease loci, they cannot detect recent mutations or those which are not widespread throughout the population. A recent GWAS on hearing loss (made available as a preprint), which reports both common and rare variant association analyses, found that the rare variant association signals were mostly independent of the common variant associations nearby, confirming that it is important to include consideration of rare variants in the genetic landscape of ARHL [10].

Alternative approaches are therefore required to identify novel variants and genes associated with age-related hearing loss. Here we have investigated variants associated with self-reported hearing difficulty in 94,312 UK Biobank participants with available exome sequence data. We have assessed variant load in self-reported hearing difficulty at a range of variant minor allele frequencies, from rare variants (minor allele frequency (MAF) < 0.005) to very common variants, and compared the resulting gene lists to a much larger list of known deafness genes that we have curated and present here, based on work in mice as well as in humans. We found the optimal MAF cutoff to be 0.1, which is an intermediate frequency, neither rare nor common. Our results suggest that intermediate frequency variants with a high predicted impact contribute to hearing difficulty, and also that the genetic contributions to hearing impairment may differ between the sexes.

## Results

After filtering, in the normal hearing group there were 18235 male (average age=62.37 years) and 30496 female (average age=62.20 years) participants (48731 people in total, overall average age 62.29). In the hearing difficulty group, there were 24237 male (average age=63.60 years) and 21344 female (average age=62.96 years) participants (45581 people in total, overall average age 63.28). It is notable that while the overall group sizes are similar, there are many more female participants than male in the normal hearing group, reflecting the better hearing that women have later in life [11], although the average age of the participants (62-63 years) is later than the average onset of menopause, after which hearing tends to decline rapidly [12]. When we plotted the distribution of each broad ethnic grouping within each category (Additional File 2: Table S1, Fig S1), we found that there were many more Black people in the normal hearing group than in the hearing loss group (especially Black female participants) (Additional File 1: Fig S1). Similar results have been noted in previous studies [13]. The distribution of other self-reported ethnicities were broadly similar across the sex-separated groups, but it is notable that the largest difference in self-reported hearing phenotype between the sexes is in the White ethnic grouping (Additional File 1: Fig S1).

### Outlier analysis of variant load

Outlier analyses of the genomic variant loads per gene in each group of people were carried out. Briefly, for each gene, the number of variants in people with hearing difficulty was compared to the number of variants in people with normal hearing using a linear regression. Each regression analysis resulted in two lists of outlier genes; those with a much higher variant load than expected (high variant load in hearing difficulty) and those with a much lower variant load than expected (which means a high variant load in normal hearing) (Fig 1. Additional File 2: Table S2). These lists were analysed to assess the effect of allele frequency and impact setting.

**Figure 1.**
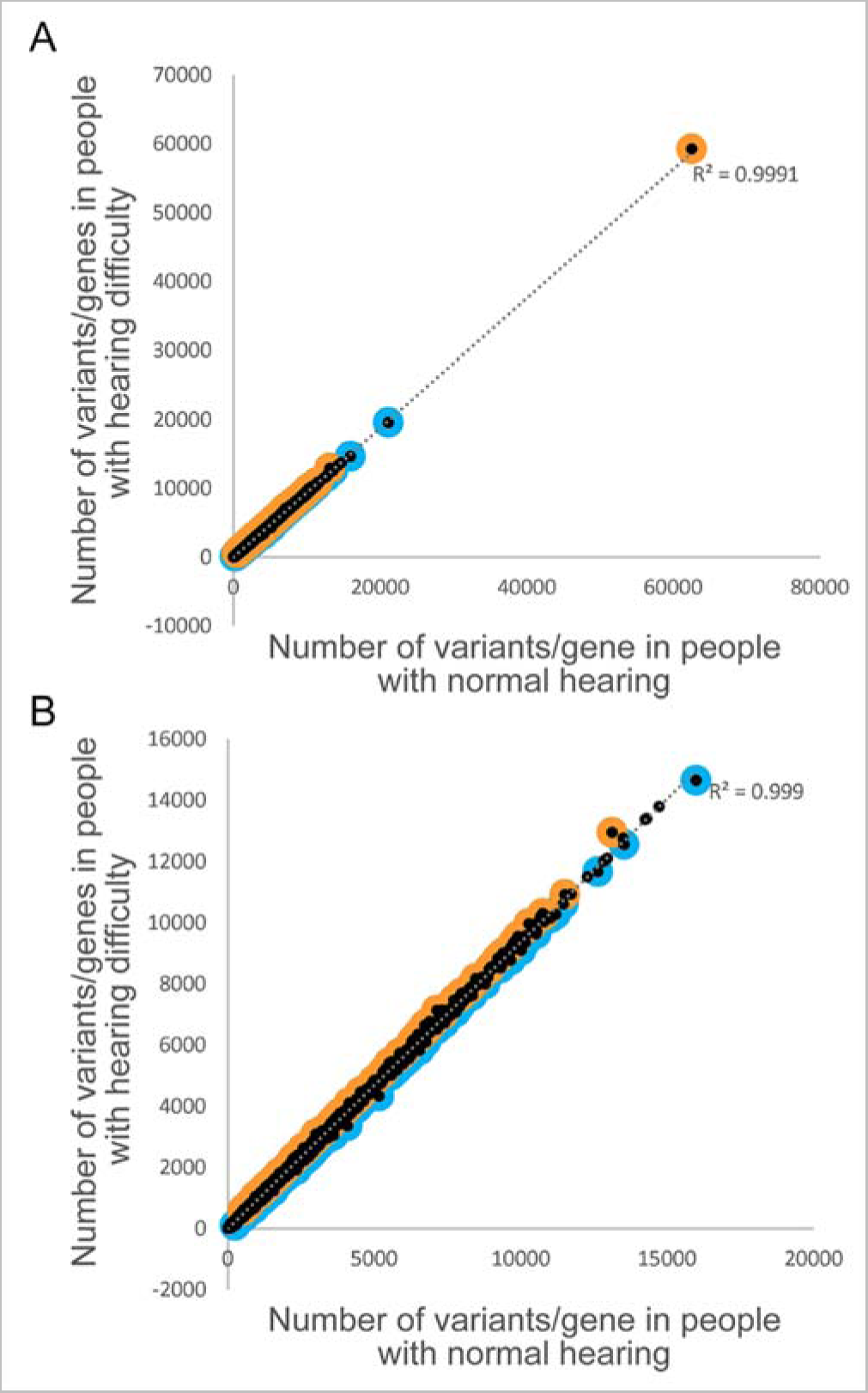
Comparison of variant load per gene for high impact variants (MAF < 0.1). Each point represents a gene. Outliers are marked in orange (for higher load in participants with hearing difficulty) or blue (for higher load in participants with normal hearing). **A** shows all the data, including *TTN* and *FBLN7*, genes with a much higher variant count than all the others, and **B** shows the data without those two genes.

We tested different minor allele frequency limits to determine the optimal cutoff. We carried out regression analyses for six MAF cutoffs (0.005, 0.01, 0.05, 0.1, 0.2 and 1), and obtained the lists of outlier genes, those genes with more variants than expected in hearing difficulty or in normal hearing (Table 1a). To assess the potential biological relevance of these high variant load gene lists to hearing impairment, genes associated with deafness in humans and/or mice were compared with genes in the two outlier lists using our own manually-curated list of known deafness genes (using the human orthologues of deafness genes known only in mice where possible, resulting in 720 genes in total) (Additional File 2: Table S3). Our assumption is that enrichment for known deafness genes supports biological relevance of the gene lists derived from the outlier analysis. We also compared the high variant load lists to our list of highly variable genes (Additional File 2: Table S4), genes which are often reported to have a high number of variants in sequencing projects. Our assumption is that enrichment for highly variable genes in the outlier gene lists is likely to reflect features unrelated to hearing. Hypergeometric tests were carried out to assess the significance of the number of deafness and highly variable genes in each outlier gene list.

**Table 1.**
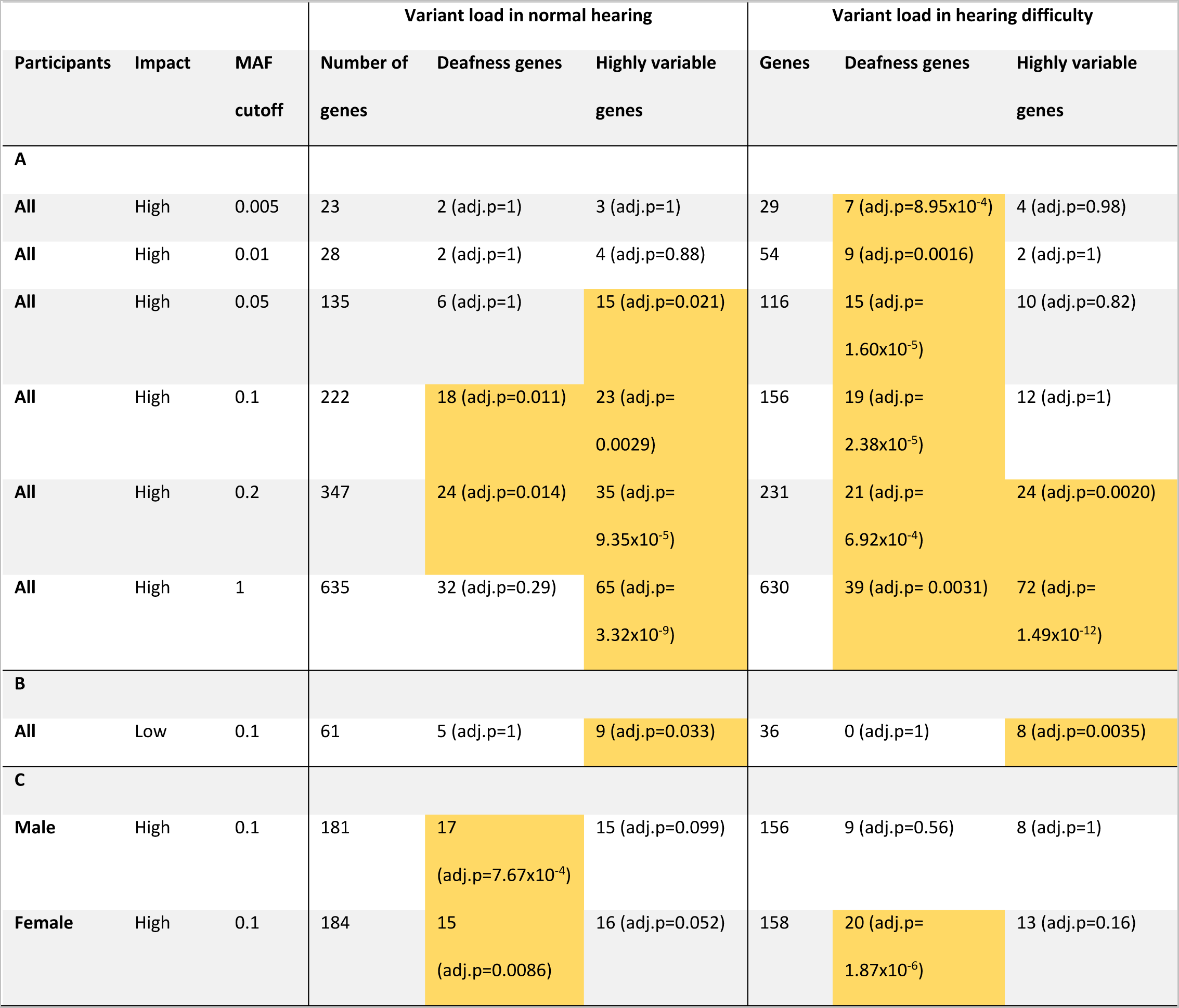
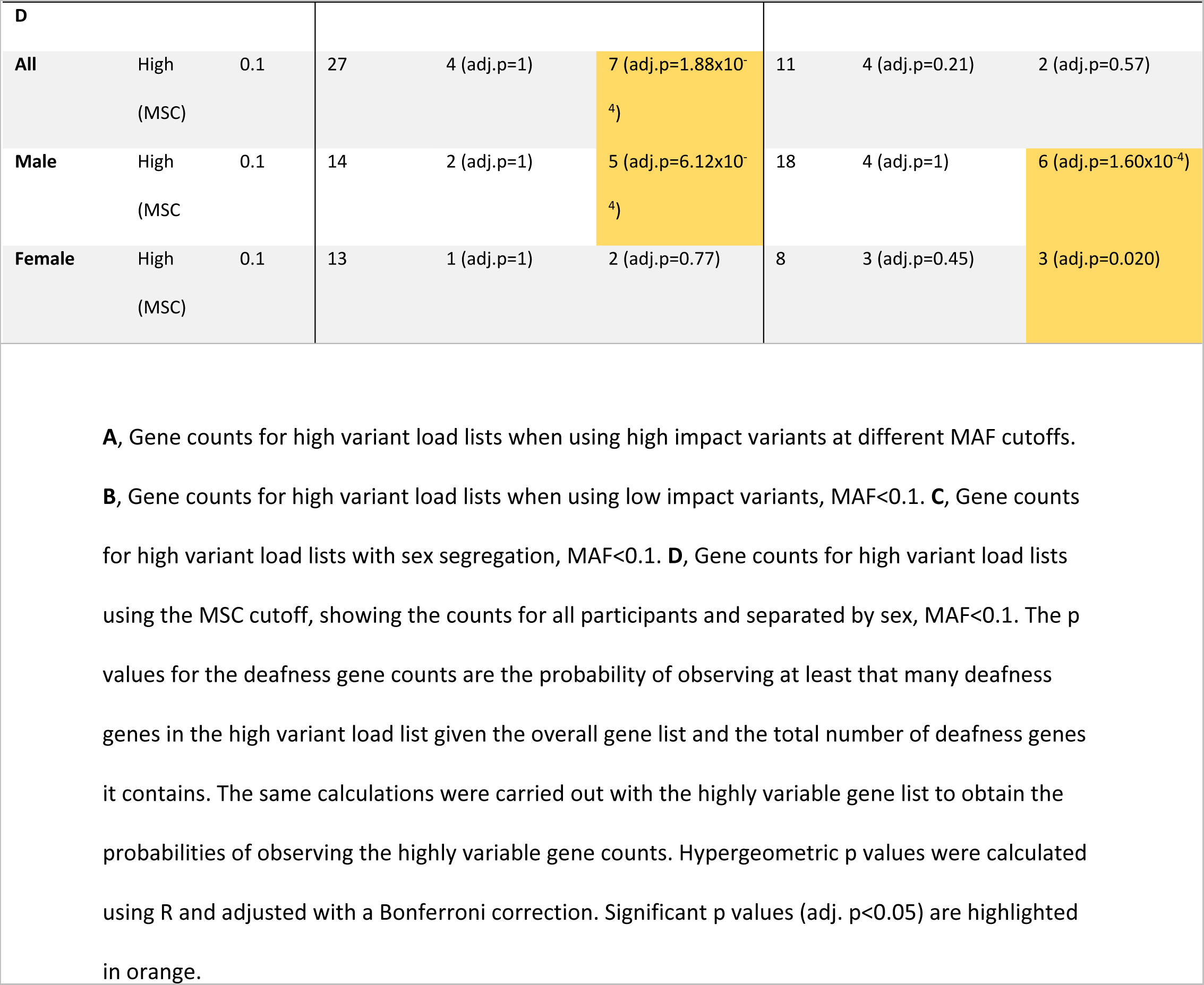
The number of genes, known deafness genes and highly variable genes in the high variant load lists at different minor allele frequencies and impacts.

|  |  |  | Variant load in normal hearing |  |  | Variant load in hearing difficulty |  |  |
| --- | --- | --- | --- | --- | --- | --- | --- | --- |
| Participants | Impact | MAF cutoff | Number of genes | Deafness genes | Highly variable genes | Genes | Deafness genes | Highly variable genes |
| A |  |  |  |  |  |  |  |  |
| All | High | 0.005 | 23 | 2 (adj.p=1) | 3 (adj.p=1) | 29 | 7 (adj.p=8.95x10<sup>-4</sup>) | 4 (adj.p=0.98) |
| All | High | 0.01 | 28 | 2 (adj.p=1) | 4 (adj.p=0.88) | 54 | 9 (adj.p=0.0016) | 2 (adj.p=1) |
| All | High | 0.05 | 135 | 6 (adj.p=1) | 15 (adj.p=0.021) | 116 | 15 (adj.p=1.60x10<sup>-5</sup>) | 10 (adj.p=0.82) |
| All | High | 0.1 | 222 | 18 (adj.p=0.011) | 23 (adj.p=0.0029) | 156 | 19 (adj.p=2.38x10<sup>-5</sup>) | 12 (adj.p=1) |
| All | High | 0.2 | 347 | 24 (adj.p=0.014) | 35 (adj.p=9.35x10<sup>-5</sup>) | 231 | 21 (adj.p=6.92x10<sup>-4</sup>) | 24 (adj.p=0.0020) |
| All | High | 1 | 635 | 32 (adj.p=0.29) | 65 (adj.p=3.32x10<sup>-9</sup>) | 630 | 39 (adj.p= 0.0031) | 72 (adj.p=1.49x10<sup>-12</sup>) |
| B |  |  |  |  |  |  |  |  |
| All | Low | 0.1 | 61 | 5 (adj.p=1) | 9 (adj.p=0.033) | 36 | 0 (adj.p=1) | 8 (adj.p=0.0035) |
| C |  |  |  |  |  |  |  |  |
| Male | High | 0.1 | 181 | 17 (adj.p=7.67x10<sup>-4</sup>) | 15 (adj.p=0.099) | 156 | 9 (adj.p=0.56) | 8 (adj.p=1) |
| Female | High | 0.1 | 184 | 15 (adj.p=0.0086) | 16 (adj.p=0.052) | 158 | 20 (adj.p=1.87x10<sup>-6</sup>) | 13 (adj.p=0.16) |

| **D** |  |  |  |  |  |  |  |  |
| --- | --- | --- | --- | --- | --- | --- | --- | --- |
| **All** | High<br>(MSC) | 0.1 | 27 | 4 (adj.p=1) | 7 (adj.p=1.88x10<sup>-4</sup>) | 11 | 4 (adj.p=0.21) | 2 (adj.p=0.57) |
| **Male** | High<br>(MSC) | 0.1 | 14 | 2 (adj.p=1) | 5 (adj.p=6.12x10<sup>-4</sup>) | 18 | 4 (adj.p=1) | 6 (adj.p=1.60x10<sup>-4</sup>) |
| **Female** | High<br>(MSC) | 0.1 | 13 | 1 (adj.p=1) | 2 (adj.p=0.77) | 8 | 3 (adj.p=0.45) | 3 (adj.p=0.020) |

**A**, Gene counts for high variant load lists when using high impact variants at different MAF cutoffs.
**B**, Gene counts for high variant load lists when using low impact variants, MAF<0.1. **C**, Gene counts for high variant load lists with sex segregation, MAF<0.1. **D**, Gene counts for high variant load lists using the MSC cutoff, showing the counts for all participants and separated by sex, MAF<0.1. The p values for the deafness gene counts are the probability of observing at least that many deafness genes in the high variant load list given the overall gene list and the total number of deafness genes it contains. The same calculations were carried out with the highly variable gene list to obtain the probabilities of observing the highly variable gene counts. Hypergeometric p values were calculated using R and adjusted with a Bonferroni correction. Significant p values (adj. p<0.05) are highlighted in orange.

As the MAF limit increased, the number of genes in the outlier lists also increased, as did the number of deafness and variable genes in each outlier list (Table 1a). However, while the number of deafness genes in the high variant load in hearing difficulty outlier list was significant at every MAF limit (Table 1a), the number of variable genes in the high variant load in hearing difficulty outlier lists did not reach significance until the MAF limit was set to 0.2 (20%). This suggests that a MAF of 0.1 is a good choice in order to obtain an outlier gene list which is relevant to hearing impairment and does not include too many highly variable genes. For the outlier genes with a high variant load in normal hearing, the MAF cutoffs where the number of deafness genes was significant (MAF < 0.1, MAF < 0.2) also resulted in a significant number of highly variable genes (Table 1a).

The effect of including variants with a low impact (Additional File 2: Table S5) was then tested, using a MAF cutoff of 0.1, and we found that we did not obtain as many genes in the outlier lists, and only the number of highly variable genes in each outlier list was significant (Table 1), suggesting that relaxing the restriction on variant impact is likely to result in detecting naturally variable genes as outliers rather than genes linked to the phenotype under study. We therefore proceeded with analysing variants with a high impact and a minor allele frequency below 0.1. From these settings, we obtained 156 outlier genes with a high variant load in hearing difficulty, and 222 outlier genes with a high variant load in normal hearing (Figure 1, Table 1a).

Because a much higher proportion of the hearing difficulty group was male, the same analysis was carried out on participants separated by sex. The numbers of outlier genes in each case were similar (Table 1b), but the gene lists were markedly different. Twenty-five genes were present in both male and female hearing difficulty high variant load lists, including seven deafness genes (*CLIC5*, *MYH14*, *COL9A3*, *ELMO3*, *FSCN2*, *GJB2*, *SLC26A5*). Twenty-four genes were present in both normal hearing high variant load lists, including three deafness genes (*POLG*, *GLI3*, *MYO3A*) (Figure 2, Additional File 2: Table S2). There was a significant enrichment in deafness genes in the normal hearing outlier lists in both sexes, and in the hearing difficulty outlier list in female participants (Table 1c). There were no significant overlaps with the highly variable gene list (Table 1c).

**Figure 2.**
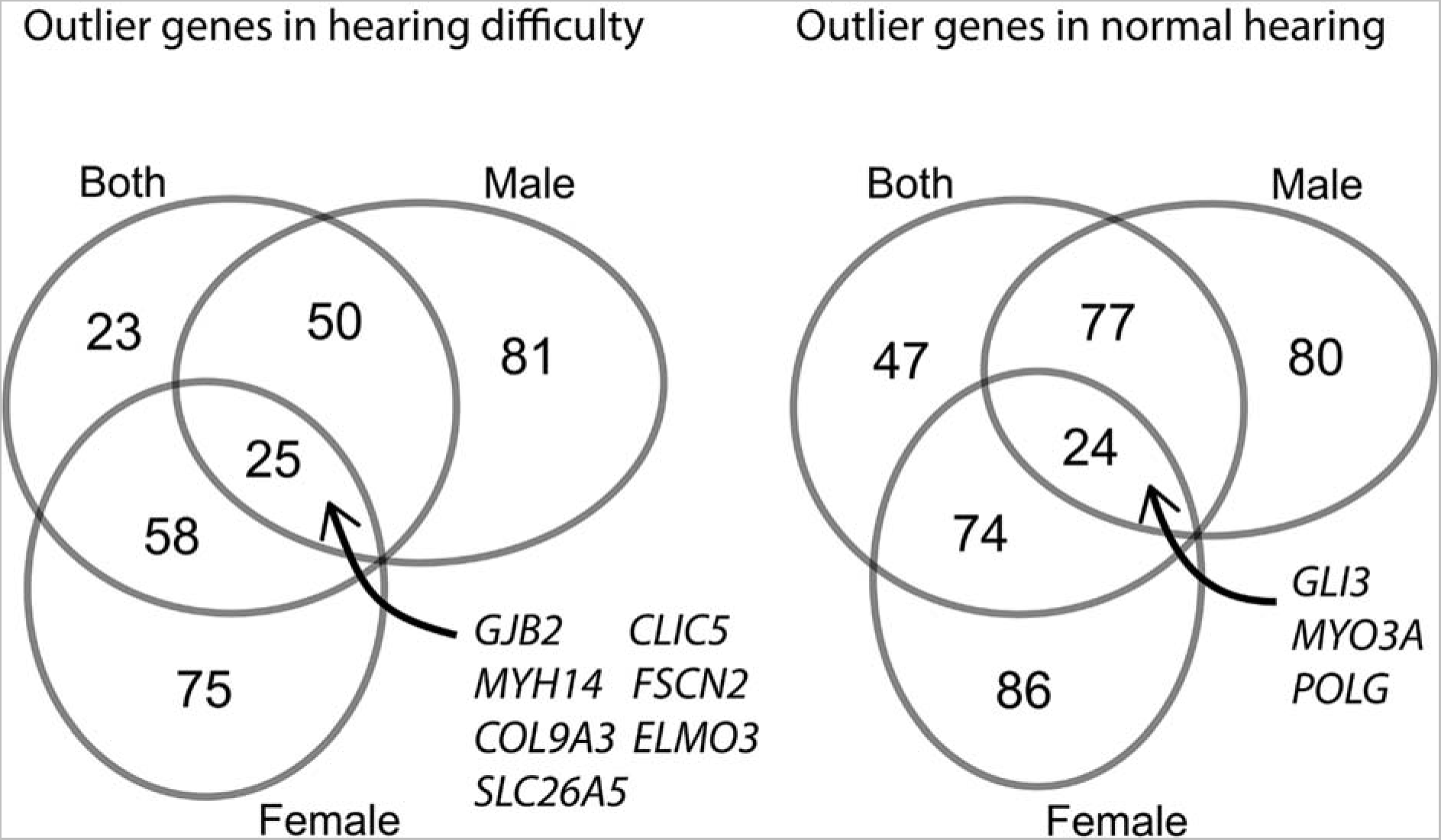
Venn diagrams showing the overlap of the outlier gene lists when looking at only male, only female, or all participants (outliers with intermediate variants with high impact). The known deafness genes in the intersection (7 in the outlier genes in hearing difficulty, 3 in the outlier genes in normal hearing) are labelled.

We chose a stringent fixed cutoff for the CADD score of 25, but it is unlikely that a single cutoff will be uniformly accurate for every gene. The mutation significance cutoff (MSC) is a gene-specific cutoff value which uses data from HGMD and ClinVar [14]. Because most genes do not have sufficient high-quality mutations described in these databases, the outlier analysis was repeated on the 2947 genes which did (MAF<0.1). We found 27 genes with a high variant load in normal hearing and 11 with a high variant load in hearing difficulty in all participants (Table 1d). Numbers in the sex-separated analyses were lower, and only the highly variable genes showed significant enrichment, in a subset of the lists (Table 1d) [15-19].

### Characteristics of the variants in the high variant load lists

The large numbers of outlier genes in people with normal hearing was unexpected, so we asked if there might be different types of variants common in hearing difficulty compared with normal hearing. We investigated the characteristics of the variants in the high variant load lists, taking the most deleterious consequence for each variant in each gene (defined in order in Additional File 2: Table S5). Variant counts were normalised per person and per gene. We did not see any large differences in variant type (Additional File 1: Fig S2). In all analyses, missense variants made up a large proportion of the total variant counts per person per gene.

### Weighted burden tests

Weighted burden analyses were carried out on the variants with MAF < 0.1 and a high predicted impact, using the geneVarAssoc and scoreassoc tools, which have been used before on the ethnically heterogenous UK Biobank dataset [20]. Scoreassoc assigns a score per subject, per gene, and tests for the difference in average scores of cases vs controls, obtaining a p value for each gene. Variants were weighted by minor allele frequency (the lower the MAF, the higher the weight), but not by impact, since all variants included in this analysis were high impact. After correcting for multiple tests, none of the genes retained significant p-values. We therefore ranked the gene list by signed log P value (SLP) [21]. The SLP is the log10 of the p-value, with a positive sign indicating that cases have more variants than controls, and a negative sign indicating the opposite. Thus, ranking the genes by SLP would result in one extreme of the list being genes with more variants in the people who did not report hearing difficulty (n=270 genes with SLP < -2), and the other extreme being genes with more variants in the people who did report difficulty hearing (n=362 genes with SLP > 2) (Additional File 2: Table S2). We carried out hypergeometric tests on these gene lists, comparing the total number of genes with the number of deafness and highly variable genes (Additional File 2: Tables S3 and S4), and found no significant enrichment of either deafness or variable genes.

### Gene ontology enrichment analysis

In order to look for any clues to pathological mechanisms, we carried out a gene ontology (GO) enrichment analysis using gProfiler [22] on the high variant load lists from the outlier analysis (Additional File 2: Table S2; high impact variants with MAF<0.1). We restricted the output to GO terms with between 5 and 200 genes, since terms with more genes than that are overly general, and those with fewer genes are too specific. We found 33 GO terms enriched in the lists, including multiple terms specific to hearing (eg GO:0007605; sensory perception of sound) (Additional File 2: Table S6). The largest list of GO terms came from the genes with a high variant load in male participants with normal hearing (Additional File 2: Table S6), mostly because of a set of genes identified as being involved in stereocilium structure and function. There were 7 genes annotated with the term “stereocilium bundle”; *USH1C*, *USH2A*, *MYO3A*, *TMC2*, *ADGRV1*, *PDZD7* and *PKHD1L1*. Most are known human deafness genes, but *PKHD1L1* and *TMC2* have only been identified as mouse deafness genes to date [23, 24]. Far fewer specific GO terms were identified from the genes with a high variant load in female participants with hearing difficulty or with normal hearing (Additional File 2: Table S6), even though there was a similar number of known deafness genes in the lists (Table 1). The term “sensory perception of sound” was identified as enriched in the hearing difficulty outlier genes in female and all participants, and in the normal hearing outlier genes in male participants (Additional File 2: Table S6). Genes annotated with this term which had a high variant load in female participants with hearing impairment were *MYO3B*, *MYH14*, *CDH23*, *CLIC5*, *CHRNA10*, *FBXO11*, *TMC1*, *GJB2*, *NAV2*, *LOXHD1*, *SLC26A5* and *MYO6*. Genes annotated with this term which had a high variant load in all participants with hearing loss were *COL11A1*, *MYH14*, *CDH23*, *CLIC5*, *CHRNA10*, *FBXO11*, *TMC1*, *GJB2*, *LOXHD1*, *SLC26A5* and *MYO6*. Most of these are known human deafness genes, but to date, *FBXO11* has only been identified in the mouse, not in humans, and *MYO3B* and

*CHRNA10* are not in our compiled list of known deafness genes (they are included in the GO term annotation through orthologous similarity rather than published evidence). In summary, the GO term analysis showed enrichment for terms relating to sensory hair cells or cytoskeletal elements known to be important to hair cell function, and this appears to be driven by the enrichment for known deafness genes in the lists analysed. Many more genes were included in the high variant load lists than were described by GO annotation terms, reflecting the limitations in current GO annotations of many genes.

### Gene prioritisation

The lists of genes of interest from the outlier analyses contained many genes not previously associated with hearing impairment, too many to follow up in detail. Therefore, ToppGene [25] was used to prioritise the genes from the high variant load lists, using our manually-curated deafness gene list (Additional File 2: Table S3) as a training list. The remaining genes in each high variant load list were scored, ranked and assigned p-values; after correction for multiple testing, we obtained eight genes from the hearing difficulty lists (*NTRK1*, *TGFBR1*, *CACNA1S*, *P2RX7*, *MYLK*, *TTN*, *CACNB3* and *ITGB1*) and three from the normal hearing lists (*NRG1*, *CACNA1H* and *FLNA*) (Additional File 2: Table S7).

### Using expression analysis to highlight new candidate genes

An analysis of the expression of candidate genes from our outlier analyses was carried out using single-cell RNAseq datasets from the gEAR database of mouse inner ear tissue analyses [26]. We reasoned that if a gene shows strong, specific expression in certain cell types in the inner ear, that suggests a potential functional role for the gene in those cell types, and would make it a good candidate for further investigation. Mouse datasets were chosen to cover as many inner ear cell types as possible between embryonic day (E) 16 and postnatal day (P) 35. We selected those candidate genes from the outlier analysis high variant load lists (Additional File 2: Table S2; high impact variants with MAF<0.1) which had a high quality one-to-one mouse orthologue (n=564), and we also plotted data for the genes known to underlie deafness in both mice and humans which were not already included (an additional 99 genes) as useful markers of cochlear cell types. After selecting those genes showing high variance in expression across the cell types, there were 234 genes from the high variant load lists and 78 more from the known deafness gene list. From the resulting heatmap, clusters of genes were annotated according to their expression, defining high expression levels (red) as 2-3, middle expression levels (orange) as 1-2 and low expression levels as 0-1 (yellow) (Additional File 1: Fig S3, Additional File 2: Table S8). Gene clusters were linked to specific cell types if they showed high or middle expression specific to those cell types, and the clusters were classified by known marker genes for specific cell types where these were present (Additional File 1: Fig S3, Additional File 2: Table S8). This allowed us to identify good candidate genes from those outlier genes which were not already known deafness genes. For example, there were three genes which appeared to be strongly and specifically expressed in pillar cells, *Col4a4* and *Col4a3*, which are known deafness genes [27], and *Thsd7a*, which is a gene with high variant loads in female and all participants with self-reported hearing difficulty (cluster 2K, Additional File 1: Fig S3). There were multiple clusters of genes strongly expressed in hair cells (clusters 1C, 1D, 1F, 1H, 1N, 1O, 1P, 1Q, 2E, 2F, 2G, 2I, 3B, 3H, Additional File 1: Fig S3), and candidate genes from this list included *Strip2* and *Brd4*, which had high variant loads in male participants reporting hearing difficulty, and *Vwa8* and *Chrna10*, which had high variant loads in female and all participants reporting hearing difficulty.

Fewer genes were observed that were strongly and specifically expressed in the spiral ganglion neurons (SGN) and strial cell types, possibly because there were fewer datasets available in the gEAR database for these cell types compared to hair cells, pillar cells and supporting cells, but cluster 2D did show consistent SGN expression (Additional File 1: FigS3). Candidate genes from the SGN cluster include the mouse deafness gene *Ercc6* [28], which has a high variant load in female participants with normal hearing, and *Abr*, which has a high variant load in all three groups with hearing difficulty. Some genes were found which clustered in strial cell types (marginal, intermediate and basal cells, clusters 1J, 1M, 3I, 4A, Additional File 1: Fig S3). All the genes plotted on the heatmap are listed with their classifications in Additional File 2: Table S8, and a summary of the clusters and their outlier genes is in Table 2. It is notable that outlier genes are found in all but two of the clusters, suggesting that there is no one cell type or cochlear location wholly or largely responsible for adult hearing difficulty in either sex (Figure 3, Table 2).

**Figure 3.**
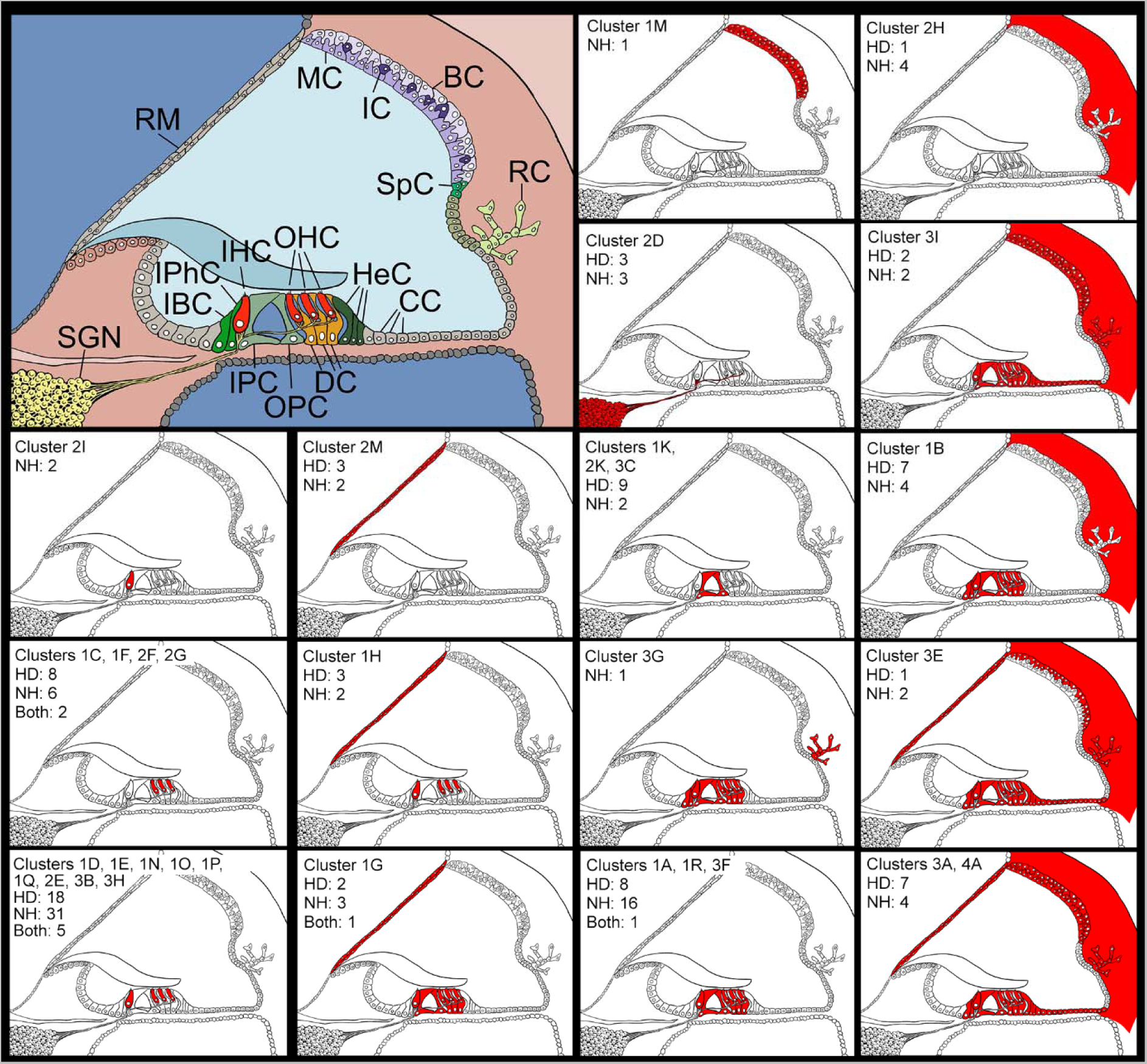
Schematic of the cochlear duct showing cell types (top left) and expression patterns based on the scRNAseq data downloaded from the gEAR database. The numbers show how many outlier genes were present in the cluster; “HD” for the number of outlier genes in hearing difficulty lists, “NH” for the number of outlier genes in normal hearing lists, and “Both” for where an outlier gene was present in a hearing difficulty list and a normal hearing list. See Table 2 for clusters and for gene names. RM=Reissner’s membrane; MC=marginal cells; IC=intermediate cells; BC=basal cells; RC=root cells; SpC = spindle cells; SGN=spiral ganglion neurons; IBC=inner border cells; IphC=inner phalangeal cell; IHC=inner hair cell; OHC=outer hair cells; HeC=Hensen cells; CC=cells of Claudius; IPC=inner pillar cell; OPC=outer pillar cell; DC=Deiters’ cells.

**Table 2.**
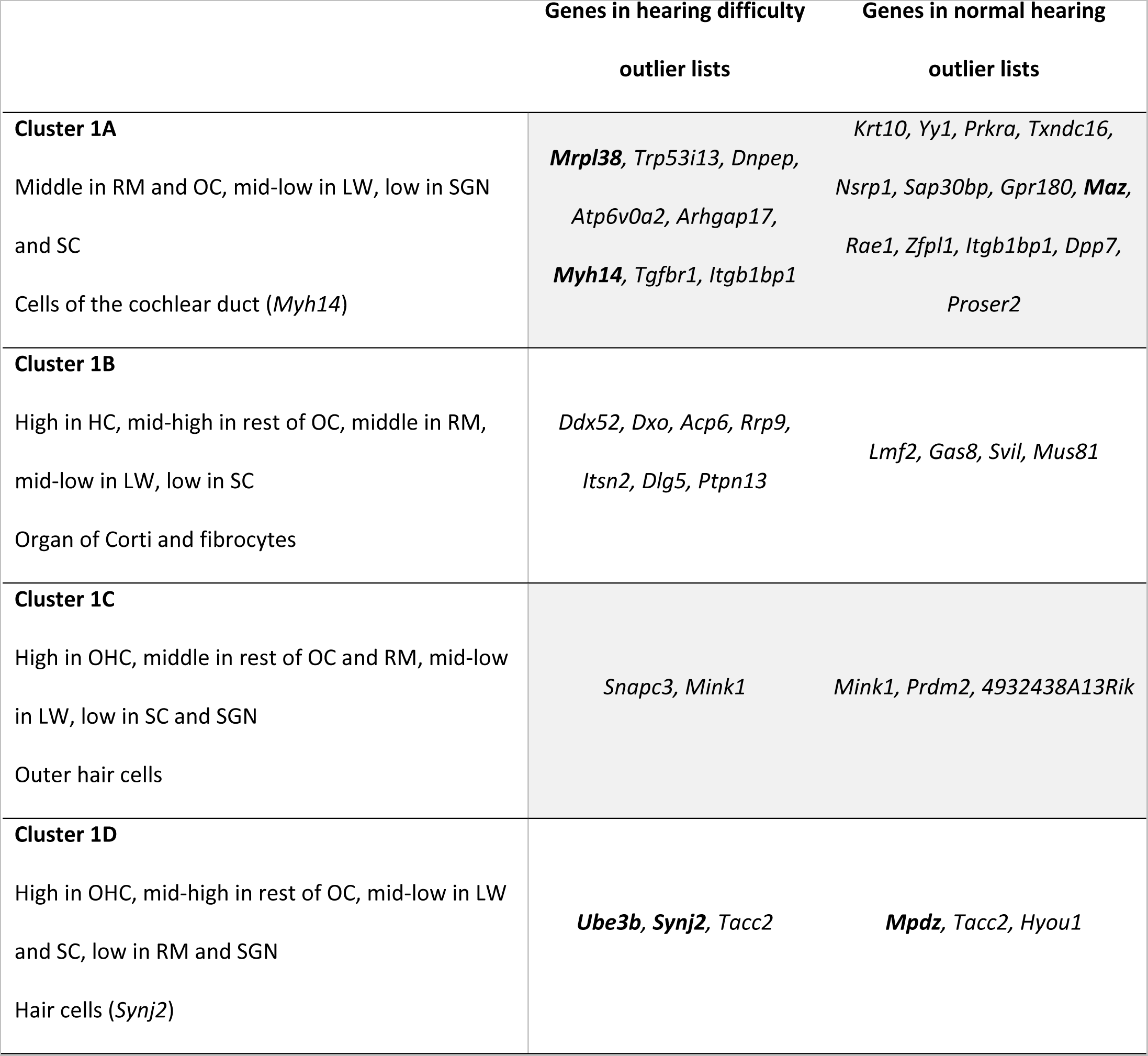
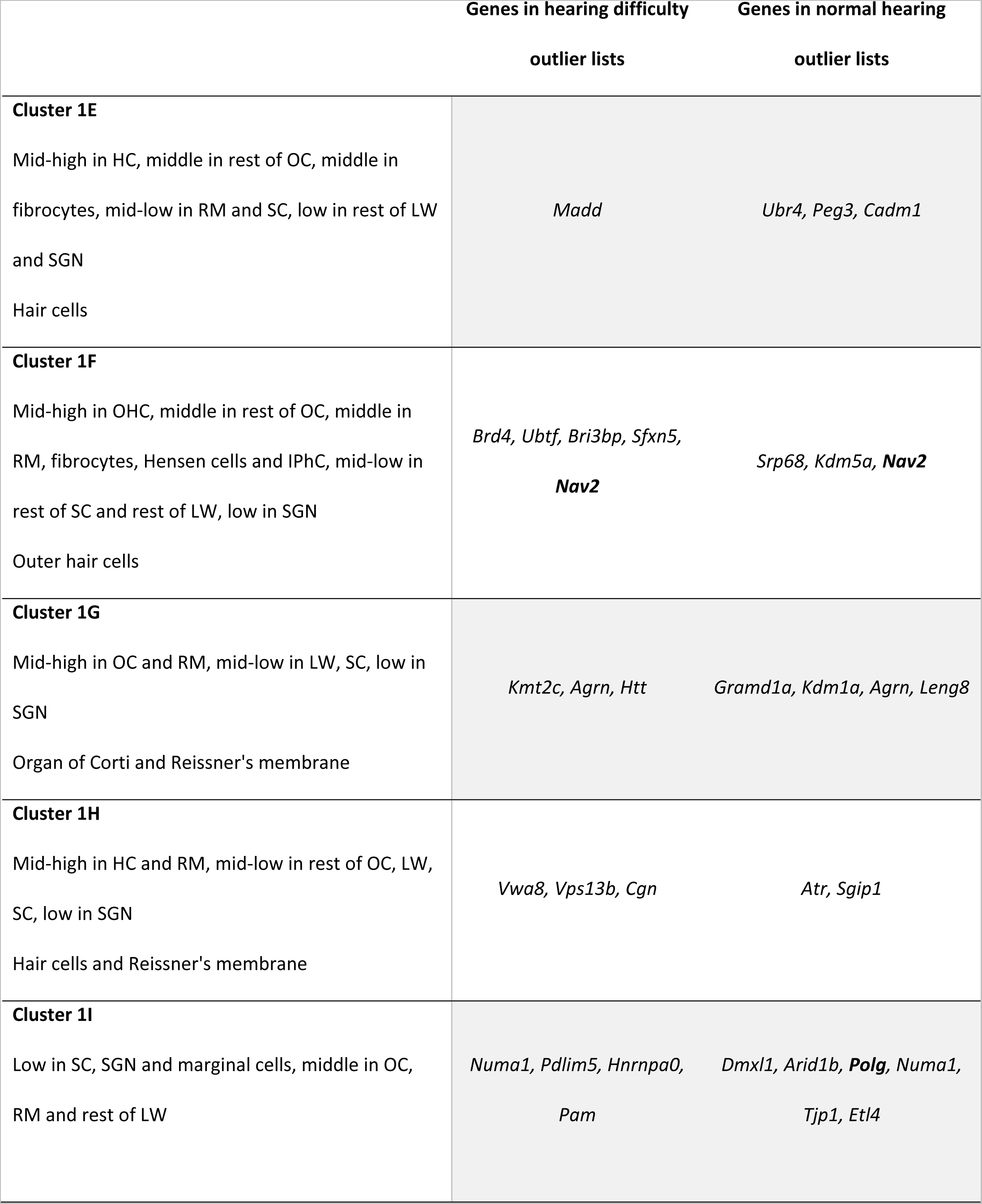
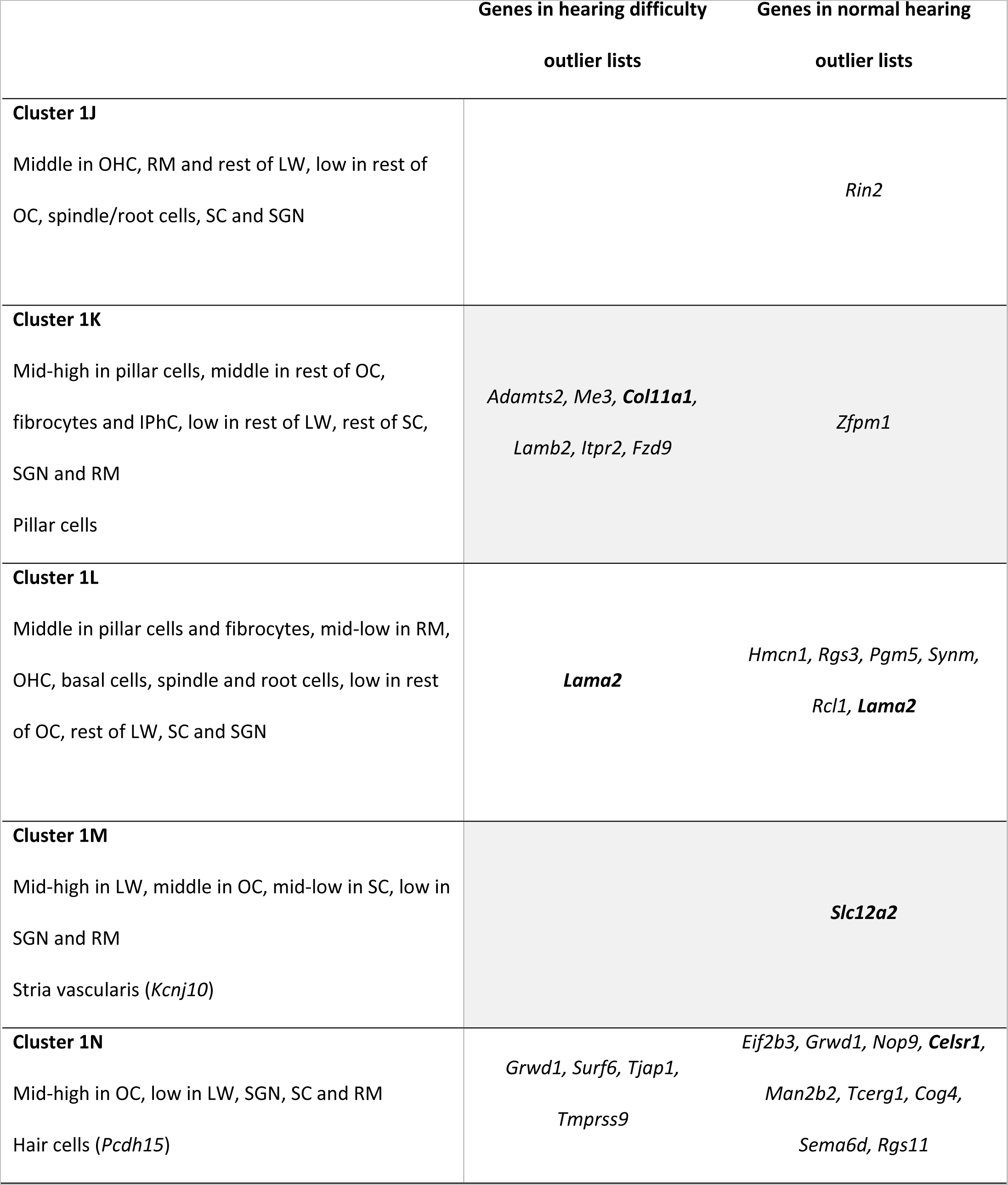
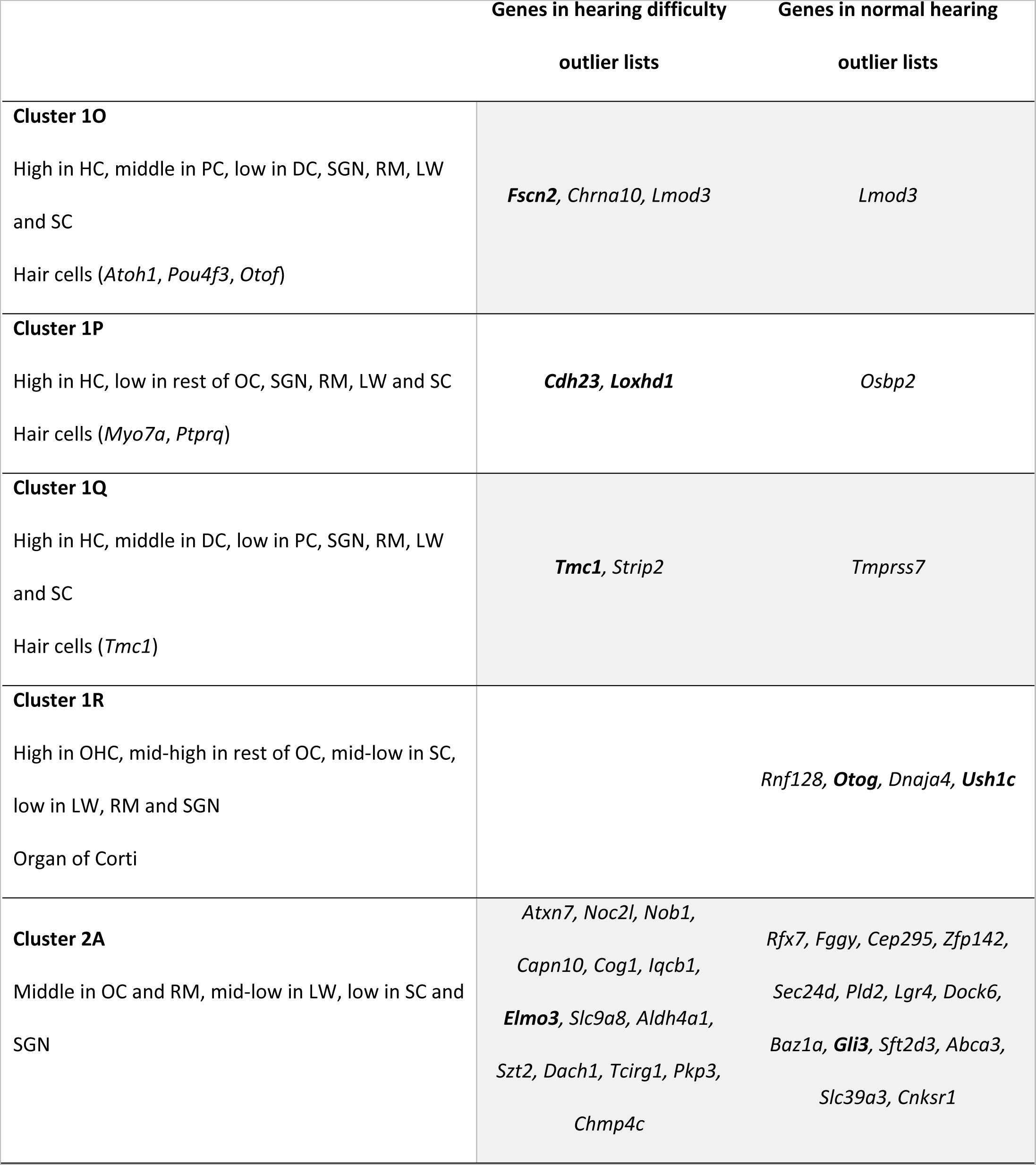
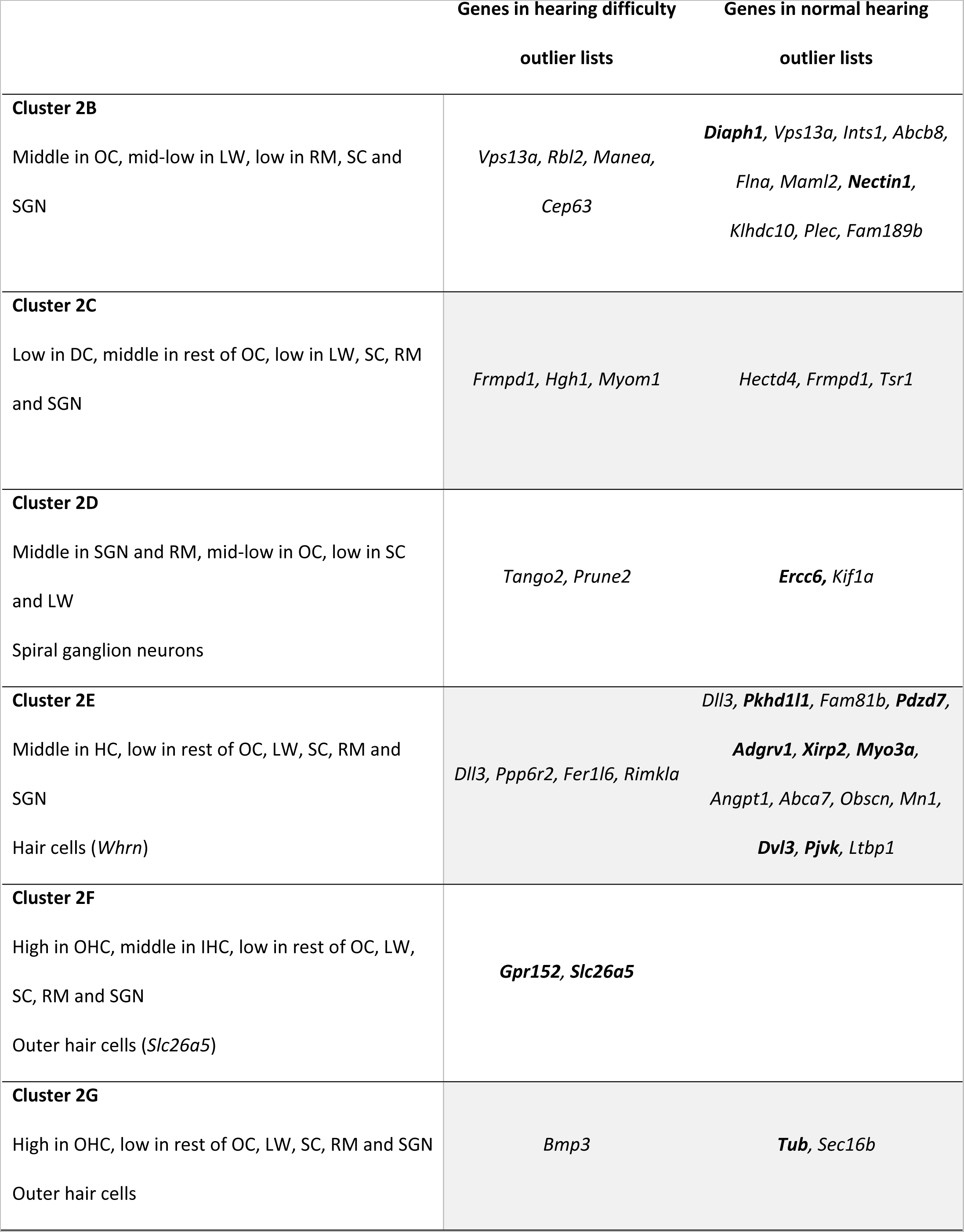
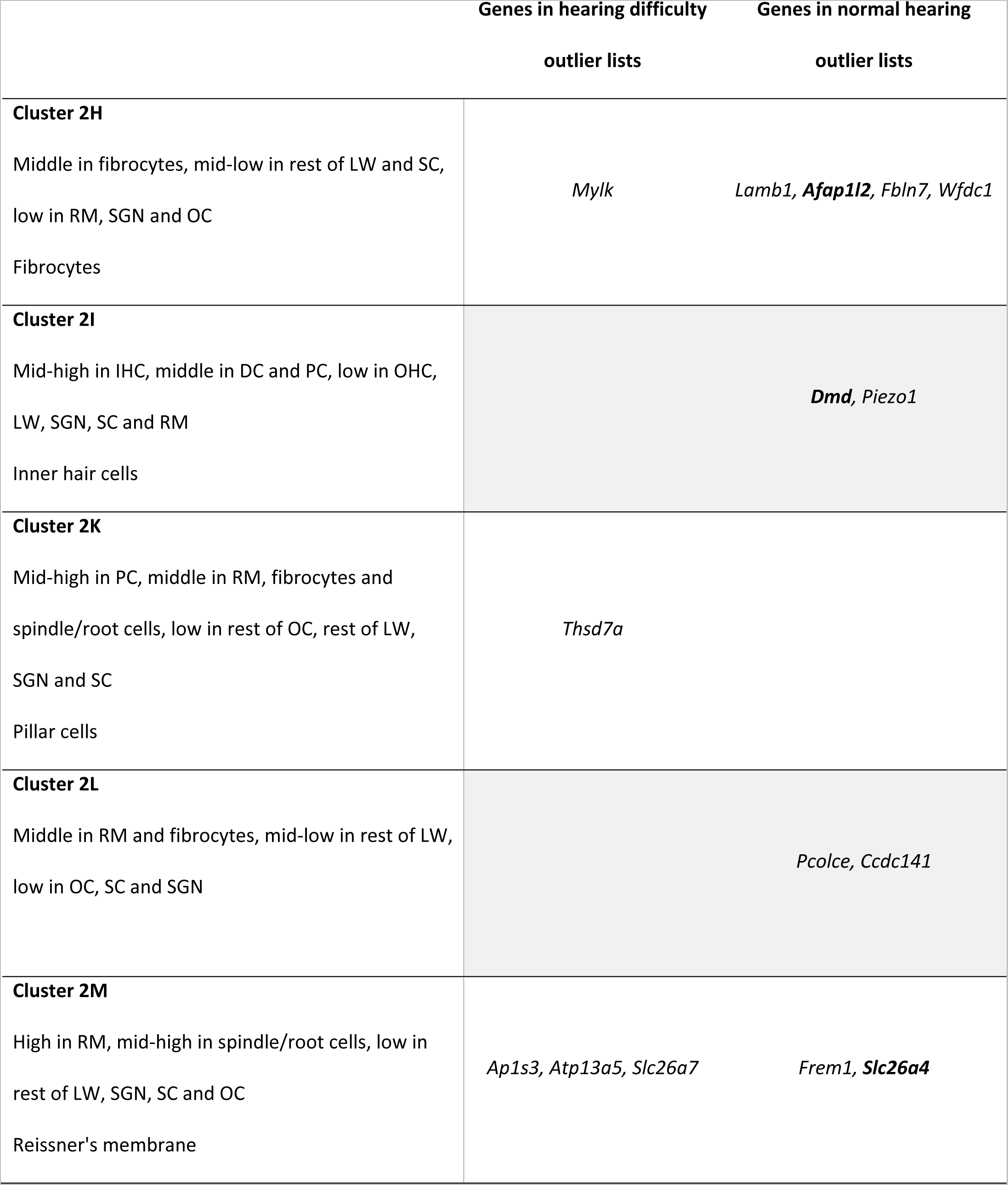
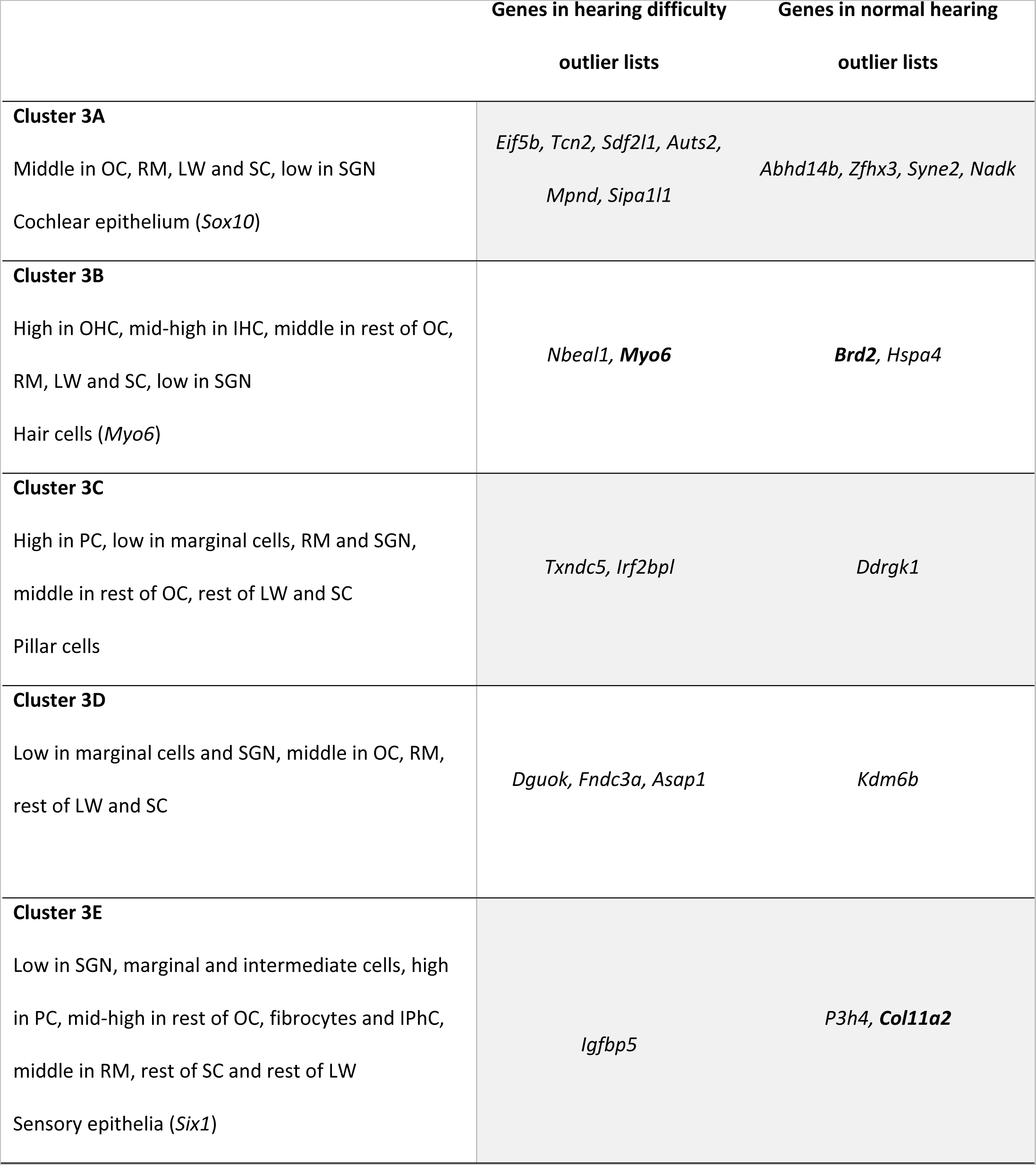
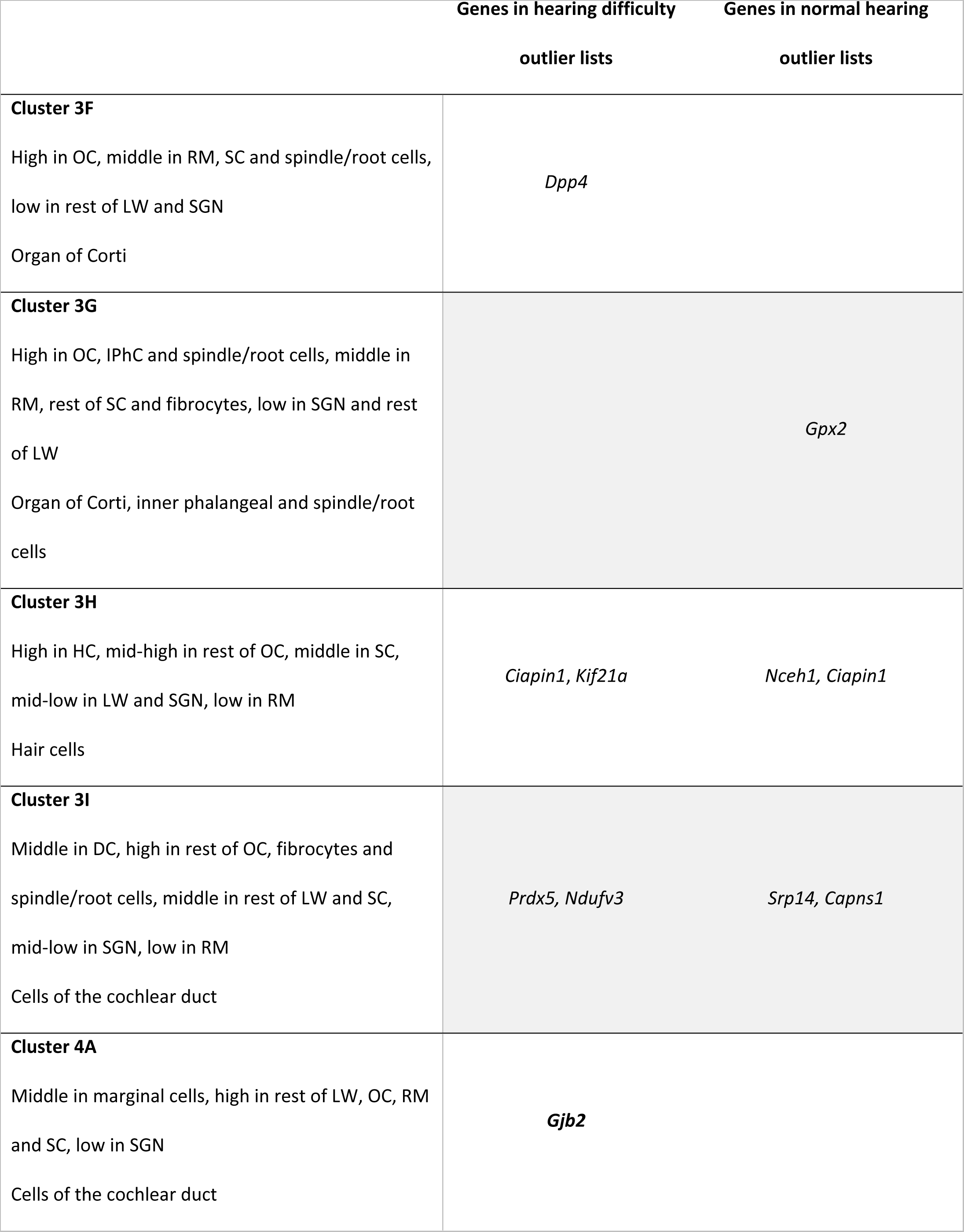
Clusters from the heatmap and their outlier genes.

All the heatmap clusters which include outlier genes are listed with their expression description, their classification (if one had been assigned) and the outlier genes separated by whether they were from hearing difficulty outlier lists or normal hearing outlier lists. Thirteen genes are present in both a hearing difficulty outlier list and a normal hearing outlier list. Known deafness genes are in bold.

## Discussion

Our data suggest that the genetic contributions to hearing difficulty in later life may differ between the sexes. That is, there may be some genetic impacts which have less effect on women than on men, and vice-versa. The differences observed in the prevalence, severity and onset of ARHL in men and women have been widely reported (for example [11, 13, 29-32], reviewed in [33]). Sex differences in complex traits and disease phenotypes may be attributed to environmental factors (in this case, noise exposure would be relevant, and drug exposure, which can affect hearing in a sex-specific manner [34]), and comorbidities which display sex-related variance may also play a role, for example cardiovascular disease [35-37]. Endogenous factors are also likely to contribute, such as hormone differences, epigenetic and regulatory differences and, of course, the different genetics involved in the XX and XY genomes. There are many studies linking estrogen to hearing sensitivity [38-40], and several genes in the estrogen pathway have been linked to hearing loss [41-43].

However, the average age of our participants was 62-63 years old, which is later than the average age of onset of menopause, so it is unlikely that hormones alone account for the observed effect. A sex-protective effect has been observed in other diseases, such that one sex requires a greater number of risk alleles to develop the disease. This was originally described by Carter et al [44, 45], who noted that women are less likely to suffer from pyloric stenosis but more likely to have children affected by the disease, but the phenomenon can apply to either sex. From our data, we found a similar number of genes bore a high load of variants in each sex, but the gene lists themselves were very different. Hearing impairment, including age-related hearing loss, while referred to as one condition, is actually the end result of a wide range of inner ear pathologies, so it is plausible that different sets of risk alleles contribute to overall hearing impairment in different sexes.

From our outlier analyses, we found the most useful MAF cutoff to be 0.1. At this level, there were 156 genes with a high variant load in people who reported hearing difficulty, and this list was significantly enriched in deafness genes but not in highly variable genes (Table 1). Increasing the MAF cutoff resulted in a significant enrichment in highly variable genes, and reducing it reduced the number of genes in the list, although the enrichment in deafness genes remained significant at all the cutoffs we tested (Table 1). This suggests that relatively common variants (MAF < 0.1) with a high predicted impact do contribute to hearing impairment, which correlates with the findings of another recent UK Biobank study [10], which reports that 16.8% of SNP heritability is contributed by “low-frequency variants” (0.001 < MAF ≤ 0.05). This is lower than our chosen cutoff (MAF < 0.1), but still higher than the standard cutoff of 0.001 recommended for autosomal dominant hearing loss [46]. Similarly, a recent report based on a very large GWAS meta-analysis also concluded that it is likely that a burden of common and rare impactful variants drives the risk of hearing loss [47]. Since ARHL is a complex disease rather than a Mendelian one, it is unsurprising that a different approach is needed when filtering for causative variants.

The burden tests we carried out did not identify any genes with a significant burden in cases (people reporting difficulty hearing) vs controls (people who did not report difficulty hearing). Burden tests compare the average of individual scores in cases and controls, while our outlier approach is an aggregated one, simply summing all the variants in cases and comparing them to the sum in controls. That has proved to be a useful approach for the UK Biobank cohort, which lacks all but the most basic auditory phenotyping data. In a cohort with more detailed auditory phenotyping, a burden test may prove to be a better approach. For example, Ivarsdottir *et al* recently reported identifying the candidate gene *AP1M2* using a loss-of-function gene-based burden test on data from a well-phenotyped Icelandic cohort [48].

Previous studies have concentrated on human hearing loss genes [2, 5], but we have compiled a larger list of nearly 700 genes based on human and mouse studies, and from this we have identified multiple candidate genes among our outliers, including *FSCN2*, *SYNJ2*, *FBXO11*, *NAV2*, *TMC2*, *ERCC6* and *PKHD1L1*. Of the 185 known human deafness genes, 118 are also mouse deafness genes (Additional File 2: Table S3), suggesting that mouse deafness genes are indeed good candidate human deafness genes. This is supported by the report from Praveen et al [10], who identified rare variant gene burdens in the mouse deafness genes *KLHDC7B*, *FSCN2* and *SYNJ2*, the latter two of which were also identified in our analyses (Table 2, Additional File 2: Table S8).

We took three approaches to explore the outlier gene lists, GO analysis, ToppGene prioritisation, and expression analysis. The GO analyses largely reiterated the comparisons with the deafness gene list (Additional File 2: Table S6). The lack of GO annotations linking the genes bearing a high variant load in hearing difficulty in the sex-separated analyses suggests that more pathways underlying hearing loss remain to be discovered and annotated. The ToppGene method is less constrained, because it uses more data sources as well as a training list to prioritise novel candidate genes, but it still relies on existing data and annotation to calculate scores and rankings. From our ToppGene prioritisation of all six high variant load lists, we obtained eleven candidate genes (Additional File 2: Table S7).

Our approach using the gEAR expression data is not limited by annotation, but is restricted to genes which have a high-quality one-to-one mouse orthologue, of which there were 564 (out of 674 outlier genes in total). It is also subject to ascertainment bias due to the relative lack of data on inner ear cell types which are not hair cells or supporting cells. We obtained 6 datasets from hair cells and supporting cells at different stages from E16 to P35, but only 2 datasets from cochlear lateral wall cell types at 2 adult ages (P20 and P30), and only one dataset from SGNs (P17-33). This means that any gene expressed during development in the lateral wall or SGNs, but not expressed in adult stages, will have been missed out of our heatmap. Additionally, most of the known deafness genes which we plotted on the heatmap are hair cell or supporting cell genes, and this may have biased the clustering. This may be why there are more outlier genes assigned to clusters with expression in hair cells (Figure 3, most notably clusters 1D, 1E, 1N, 1O, 1P, 1Q, 2E, 3B and 3H). Despite that, we did observe several clusters with expression in the lateral wall and spiral ganglion (Figure 3, Additional File 1: Fig S3). We have identified multiple potential candidate genes based on their presence in the outlier gene lists and their expression in specific cell types within the cochlea (Table 2), such as *THSD7A*, which is expressed in pillar cells, and *PRUNE2*, a gene with expression in the spiral ganglion neurons, both of which have a high variant load in hearing difficulty (female and all participants). Three of the ToppGene candidates were included in the gEAR heatmap; *TGFBR1* and *FLNA*, which are mainly expressed in the organ of Corti, and *MYLK*, which is expressed in fibrocytes (Additional File 2: Table S8).

The nature of the regression analysis means that we detected outlier genes associated with normal hearing as well as with hearing impairment, and our enrichment analyses of the outlier gene lists (high impact variants with MAF < 0.1) showed almost all of them were significantly enriched in deafness genes (outliers with a high variant load in male participants with hearing difficulty was the only exception) (Table 1). This suggests that the high variant loads are driven by the association with the self-reported hearing phenotype, not just statistical noise, sequencing error and the natural genetic variability observed in some genes, particularly large genes like *TTN* and *USH2A*. This includes the high variant loads associated with normal hearing as well as those associated with hearing impairment. It is possible that there may be protective variants in some of these genes, for example, variants which result in protection against noise trauma or ototoxic drug exposure, or which simply improve the maintenance of the inner ear machinery. Such a variant has recently been reported in *GJB6* in mice; homozygotes for the deleterious Ala88Val mutation displayed better hearing at older ages, better neural output from the inner ear, and reduced hair cell loss [49]. This is not the only precedent for deleterious mutations having a beneficial impact on a phenotype, and such mutations may be attractive targets for drug development. For example, Akbari et al (2021) recently reported that multiple rare protein-truncating variants in the gene *GPR75* were associated with protection from obesity, and mice lacking the orthologue, *Gpr75*, were resistant to weight gain on a high fat diet [50]. Similarly, rare deleterious variants in *B4GALT1* have been linked to decreased coronary artery disease via reduction of fibrinogen and low-density lipoprotein cholesterol [51]. Further investigation of the genes and variants linked to normal hearing and sex differences in hearing loss is needed.

Previously-reported GWAS of the UK Biobank which also used the self-reported hearing phenotype identified multiple overlapping loci; 71 in total, 19 of which were shared between all three studies (Figure 4) [8, 9, 48]. Four of those 19 were also identified in our study (*CHMP4C*, *NID2*, *SYNJ2* and *CDH23*). Five further genes from our outlier lists were shared between a subset of the GWAS lists; *TMPRSS9*, *LOXHD1*, and *TUB* were identified by Ivarsdottir *et al* and Kalra *et al*, and *SLC26A5* and *FSCN2* by Ivarsdottir *et al* alone. Those genes identified by multiple studies are obvious candidates for involvement in ARHL, but the differences in loci identified using the GWAS approach suggest there are many more to investigate, and the results of our exome sequencing analysis support that as well as suggesting further candidates (Table 2). It has recently been observed that rare variants do not account for the GWAS hits of common markers [10], so it is unsurprising to find different variants and different genes associated with ARHL in GWAS compared with exome/genome sequence analysis studies.

**Figure 4.**
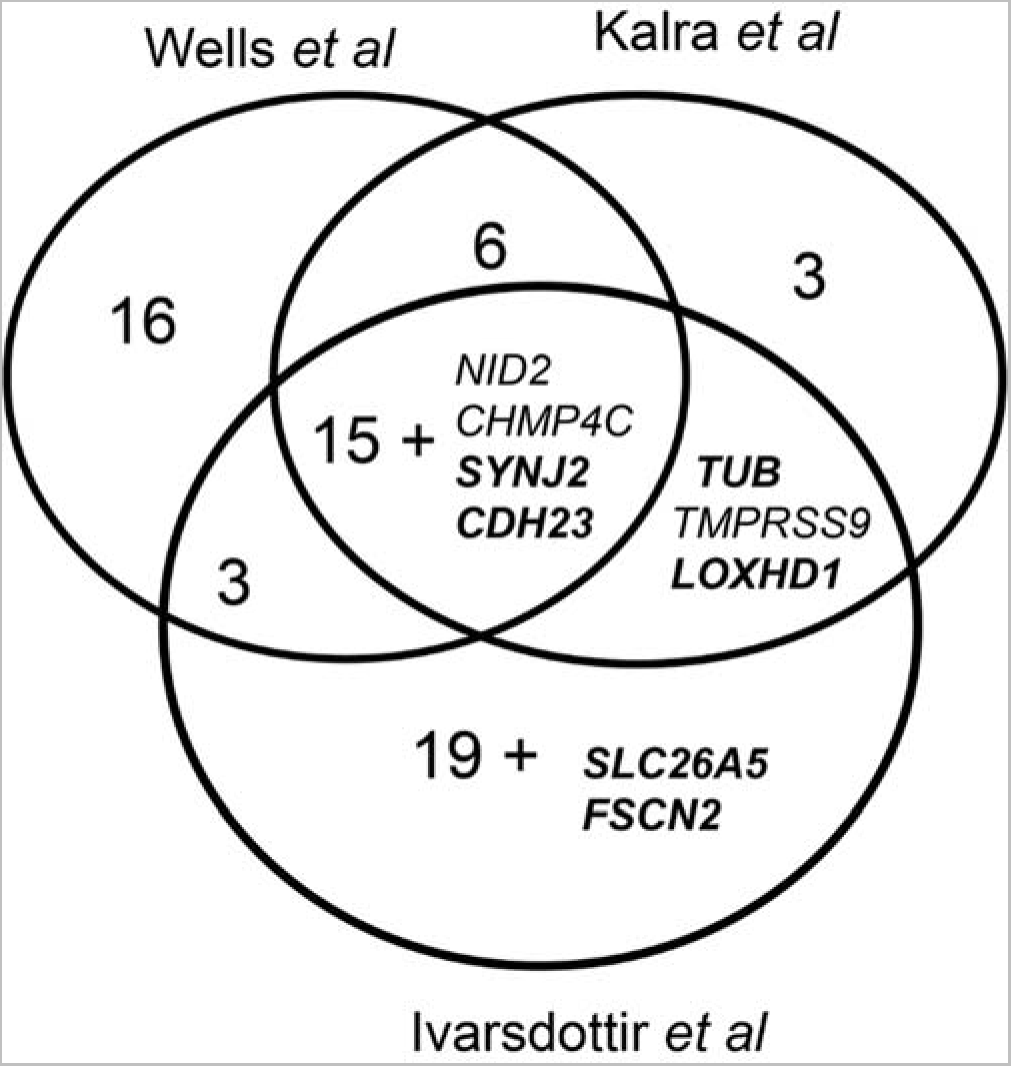
Comparison of gene lists from recent UK Biobank GWAS on self-reported hearing. Labelled genes are those also identified in this study, and are not included in the numbers for those sections. Known deafness genes are in bold. The Wells *et al* [9] and Kalra *et al* [8] analyses used the UK Biobank data only while the Ivarsdottir *et al* analysis [48] included other populations from Iceland.

The biggest limitation of this study is the lack of measured hearing impairment (such as an audiogram) and detailed auditory phenotyping. Self-reported hearing difficulty has been shown to be sufficiently informative for general hearing capacity [9, 52], but there is more to hearing loss than just an average threshold shift. It is likely that we have missed mild or even moderate hearing loss, and also unilateral hearing loss. Most notably, we were not able to exclude participants who had experienced hearing impairment from a young age (with the exception of cochlear implant users, who were excluded). Being able to compare specific subtypes of true age-related hearing loss (for example, using a classification system such as the one described in [53], [54], and [55]), offers the potential to link genes with a high variant load to specific inner ear pathologies, an important step for stratifying patient populations and developing therapeutics.

## Conclusions

From this study, we have established that it is useful to include more common variants when investigating a heterogeneous disease such as adult hearing difficulty. In this case, we found the most useful MAF cutoff to be 0.1, but it is likely that this varies by condition. We also found that the genetic contributions to self-reported hearing difficulty differ between the sexes, suggesting that in future studies, it would be useful to separate study participants by sex, as well as analysing all participants together. Future studies would also benefit from more detailed auditory phenotyping data.

While these points are based on a study of adult self-reported hearing difficulties, it is likely that they apply to many other conditions. As the availability of large-scale exome and genome sequencing studies grows, it is important to explore questions which could not be asked using earlier paradigms such as genome-wide association studies. This work highlights several such avenues of exploration.

## Methods

### UK Biobank participant selection

UK Biobank is a large-scale biomedical database and research resource containing genetic, lifestyle and health information from half a million UK participants, aged between 40-69 years in 2006-2010, who were recruited from across the UK. Participants have consented to provide their data to approved researchers who are undertaking health-related research that is in the public interest. Participants were selected who were ≥55 years of age who had exome sequencing data available (200,619 exomes available in total, September 2020) and could be classified as having normal hearing or hearing difficulty, based on their self-report of hearing difficulty, hearing difficulty in noise, or use of a hearing aid. If people reported no hearing difficulties or hearing aid use at any assessment and had been asked about their hearing at least once when they were ≥55, we included them in the “normal hearing” group. If people reported consistent or worsening hearing impairment, or that they had at any point been a hearing aid user and had been asked at least once about their hearing when they were ≥55, we included them in the “hearing difficulty” group. Participants who reported otologic disorders (eg Meniere’s disease) were excluded. People who reported high levels of noise exposure or moderate/severe tinnitus were also excluded from the normal hearing group (Additional File 1: Figure S4). This resulted in a total of 48731 people in the normal hearing group (18235 male and 30496 female participants), and 45581 people in the hearing loss group (24237 male and 21344 female participants). Overall, 106307 participants with exomes were excluded based on the above criteria. We did not filter by self-reported ethnicity. The vast majority (96%) of participants described themselves as “British”, “Irish”, “White”, or “any other White background”, or some combination thereof (hereafter referred to as White). In most of the broad ethnic groupings (Additional File 2: Table S1), there were more female than male volunteers (Additional File 1: Figure S1). Participants included in this study were compared to the entire UK Biobank, to the UK Biobank participants who had had exome sequencing, and to the data from the UK census 2011 [56], and we found that while the proportion of self-reported minority ethnicities was smaller in the UK Biobank than in the 2011 census [56], it was smaller still in the participants included in this study (Additional File 1: Fig S1). However, the distribution of self-reported ethnicities in the participants with exome sequencing reflected that of the entire Biobank (Additional File 1: Figure S1). The “healthy volunteer” effect, meaning that participants tend to be healthier in terms of lifestyle and health conditions, has been previously noted in the UK Biobank when compared to the UK 2011 census data, as has the greater proportion of people reporting their ethnicity as White [56]. It is not clear why the subset of the UK Biobank selected for this study, on the basis of their answers to questions about hearing and related issues, has an even greater proportion of participants who report their ethnicity as White.

### Variant annotation and filtering

UK Biobank variant calls were made available following processing, variant calling and joint genotyping [57, 58], but without any filters applied at the sample or variant level. We annotated the variants using the Ensembl Variant Effect Predictor [59], including data from ReMM, which provides a measure of pathogenicity for regulatory variants [60], SpliceAI, which scores variants based on their predicted effect on splicing [61], Sutr, which provides annotations for 5’UTR variants, including a predicted effect on translation efficiency [62], and the deleteriousness predictor CADD [63]. Minor allele frequencies were obtained from gnomAD (African, admixed American, Ashkenazi Jewish, East Asian, Finnish, Non-Finnish European, Other) [64], the 1000 Genomes project (African, admixed American, East Asian, European, South Asian) [65], TopMed (not divided by population) [66] or ESP6500 (African, European) [67], and the maximum reported minor allele frequency (MAF) was used. Variants were then filtered based on the overall quality of the variant call (QUAL, minimum 20) and the read depth (DP, minimum 10) and genotype quality (GQ, minimum 10) of individual calls. Variants with more than 10% of calls missing were also excluded, as were those which had a high private allele frequency within the UKBB cohort (defined as the recorded minor allele frequency + 0.4) [68]. In order to exclude variants exhibiting excess heterozygosity, we excluded variants which failed the Hardy-Weinberg equilibrium test and which had excess heterozygosity >0.1 (excess heterozygosity was calculated by (O-E)/E, where O is the observed heterozygote count and E is the expected heterozygote count).

### Variant classification filters

Variants were filtered based on their minor allele frequency and a combination of pathogenicity and consequence filters (Table 3). We defined two levels of impact upon a gene product, low impact and high impact. High impact variants were those in coding regions, intronic splice sites or mature miRNAs with a CADD score > 25 or a SpliceAI score > 0.5, and those in 5’UTRs with a Sutr score > 1. Low impact variants were all those in coding regions, intronic splice sites, mature miRNAs and 5’UTR regions, those in 3’UTR regions, and any variants with other classifications (eg regulatory region variants) which had a ReMM score > 0.95 (see Additional File 2: Table S5 for the exact variant classification terms and filters). This is an inclusive classification, so the list of low impact variants includes the high impact variants.

**Table 3.** Classification criteria for variants by impact and minor allele frequency.

|  | Consequence | Pathogenicity | Minor allele frequency | Number of variants |
| --- | --- | --- | --- | --- |
| **Meaning** | The effect of the mutation on the protein | How likely the mutation is to impair protein function | How rare the alternative allele is in the population |  |
| **Source** | Ensembl, ReMM | CADD, Sutr, SpliceAI | gnomAD, 1000G, TOPMed, ESP6500 |  |
| **High impact** | 5'UTR variants, splice site mutation, stop gain or loss, start loss, insertion, deletion, duplication, missense variants, and variants in mature miRNAs | CADD > 25 or Sutr > 1 (for 5'UTR variants only) or SpliceAI score > 0.5 (for splice site variants only) | <0.005 | 1141302 |
|  |  |  | <0.01 | 1151111 |
|  |  |  | <0.05 | 1160767 |
|  |  |  | <0.1 | 1162399 |
|  |  |  | <0.2 | 1163464 |
|  |  |  | <1 | 1165167 |
| **Low impact** | All consequences except for intergenic and intronic variants, variants in transcripts subject to nonsense-mediated decay, and variants up- or downstream of a gene; All variants with a ReMM score > 0.95 | Not assessed | <0.1 | 6398787 |

### Regression outlier analysis

For each analysis, we assessed 21841 protein-coding genes and microRNAs (Additional File 2: Table S9). We summed the total number of variants in each gene in participants from the normal hearing group and compared them to the total number of variants in that gene from the hearing difficulty group using a linear regression. The residuals were obtained for each gene (the difference between the observed and predicted variant load in hearing impairment) and the first (Q1) and third (Q3) quartiles, and the interquartile distance (D, Q3-Q1), were calculated. Outlier genes with a high variant load in hearing difficulty were defined as those with residuals > Q3 + 6D, and outlier genes with a high variant load in normal hearing were defined as those with residuals < Q1 – 6D [69]. All participants were subjected to these comparisons, and we also carried out sex-separated analyses. Hypergeometric distribution tests were carried out using R, and gProfiler [22] was used to carry out a GO enrichment analysis of the outlier gene lists. The Bonferroni correction was used to adjust for multiple testing.

### Burden tests

Weighted burden tests were carried out using the geneVarAssoc/scoreassoc software [21, 70], which has been shown to be capable of handling heterogeneous datasets [20]. Population principal components were derived from common variants using plink v2.0, following reading in of variants from vcf files using plink v1.9 [71]. For each gene, scoreassoc assigns scores to subjects according to the variants carried, assigning weights according to minor allele frequency, such that rarer variants are assigned a higher weight. The software then tests whether the average score for cases is higher than the score for controls [21]. The Bonferroni correction was used to adjust for multiple testing.

### Compilation of the list of deafness genes

We compiled a manually curated list of known deafness genes in humans and mice, including all genes listed in the Hereditary Hearing Loss Homepage [1], and genes which, when mutated, result in altered hearing thresholds in mutant mice, reported by the International Mouse Phenotyping Consortium (www.mousephenotype.org [72, 73]; average thresholds were individually checked for shifts >10dB with small standard deviations). We also included mouse and human deafness genes described in the literature (for example [74, 75]; for full reference list see Additional File 2: Table S3). There were 118 genes shown to underlie hearing in mice and humans, 67 human deafness genes (with 66 mouse orthologues) and 506 mouse deafness genes (with 535 human orthologues) (Figure 5, Additional File 2: Table S3). Although many of these known deafness genes have only been linked to early-onset, severe hearing impairment, they are still good candidates for involvement in milder hearing impairment, since different variants can result in very different phenotypes. For example, different variants in *TMC1* have been shown to result in either prelingual profound hearing loss or postlingual progressive hearing loss [76, 77], and several recent large-scale studies looking at adult-onset hearing loss have found multiple missense variants in Mendelian deafness genes with milder effects than previously reported [5, 10, 48].

**Figure 5.**
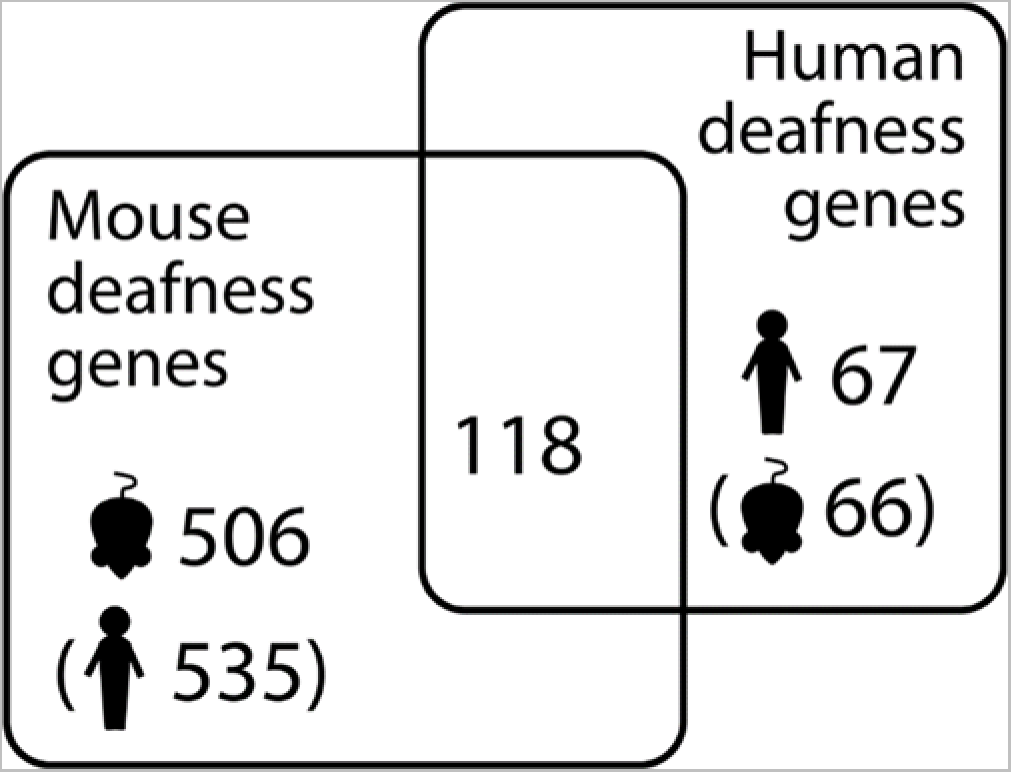
Deafness gene counts in mice and humans. Brackets indicate orthologues (eg there are 66 mouse orthologues of the 67 human deafness genes).

### Compilation of the list of highly variable genes

Some genes are often reported as having a high number of variants in multiple exome sequencing projects. This can be because they are large genes (eg *TTN*), or because they belong to groups of paralogues such as olfactory receptors, which are sufficiently similar to make correct alignment difficult, resulting in incorrect variant calls. Two such lists were compiled by Adams et al [78], and Fuentes Fajardo et al, [79], and consist of genes which contributed many variant calls to multiple exomes as well as human leukocyte antigen (HLA), taste receptor (TAS), olfactory receptor and mucin family genes. Additionally, some genes have been identified as prone to recurrent false positive calls, which are variants that did not validate with further genotyping and were not heritable [80]. We combined all three lists, resulting in 1213 genes in total (Additional File 2: Table S4).

### Gene expression analysis using the gEAR

To assess the expression of lists of genes of interest in the inner ear, including the list of known mouse and human deafness genes, we used single cell RNAseq data from the mouse inner ear, accessed via the gEAR portal (https://umgear.org/) [26]. We chose datasets from mice aged between embryonic day (E) 16 and postnatal day (P) 35. The datasets we used came from E16 cochlea, P1 cochlea, P7 cochlea [81, 82], P15 cochlea [83, 84], P20 inner ear [85, 86], P28-35 cochlea [87-89], P30 stria vascularis [90, 91] and P17-33 spiral ganglion neurons [92, 93][92]. Expression levels were normalised to *Hprt* expression; where *Hprt* was not present in the dataset, or had an expression level of 0, we did not use the data. We then summarised the data, taking the maximum level per cell type without accounting for age. Because the expression levels ranged from 0 to 70.6 (*Ceacam16*, in outer hair cells), we transformed the data such that levels between 10 and 100 were scaled to between 2 and 3, and levels between 1 and 10 were scaled to between 1 and 2. We used R to plot a heatmap of the genes that showed the most variability between cell types, suggestive of specific expression patterns rather than non-specific expression (n=312, variance across datasets > 0.15), and to cluster cell types and genes. We further defined gene clusters first based on the R dendrograms and then on the gene expression levels within specific cell types or groups of cell types.

## List of abbreviations

ARHL: age-related hearing loss
MAF: minor allele frequency
GO: gene ontology
GWAS: genome-wide association study

## Declarations

### Ethics approval and consent to participate

UK Biobank has ethics approval from the North West Multi-centre Research Ethics Committee (MREC), as a Research Tissue Bank (RTB) approval (number 21/NW/0157). Informed consent was obtained from all participants. Use of the relevant data for this study has been approved by the UK Biobank (ID 49593).

### Consent for publication

Not applicable.

### Availability of data and materials

All UK Biobank data, including the exome sequence data and questionnaire data used in this study, are publicly available to registered researchers through the UK Biobank data access protocol. Further information about registration may be found at http://www.ukbiobank.ac.uk/register-apply/. The mouse single cell RNAseq data used in this study are publicly available from the gEAR database at https://umgear.org, and also from the GEO repository (https://identifiers.org/geo:GSE181454, https://identifiers.org/geo:GSE114157, https://identifiers.org/geo:GSE136196, https://identifiers.org/geo:GSE137299, https://identifiers.org/geo:GSE117055, https://identifiers.org/geo:GSE111347, https://identifiers.org/geo: GSE1113478) [81, 84, 86, 88-90, 93].

### Authors’ contributions

MAL carried out the data processing, outlier analysis, burden analysis and gene expression analysis. MAL and KPS wrote the paper. All authors read and approved the final manuscript.

## Data Availability

All UK Biobank data, including the exome sequence data and questionnaire data used in this study, is publicly available to registered researchers through the UK Biobank data access protocol. Further information about registration may be found at http://www.ukbiobank.ac.uk/register-apply/. The mouse single cell RNAseq data used in this study is publicly available from the gEAR database at https://umgear.org.

https://umgear.org

## Acknowledgements

This research has been conducted using data from UK Biobank, a major biomedical database (www.ukbiobank.ac.uk, project ID 49593). The authors are grateful to everyone involved in the UK Biobank, especially the participants, without whom there would be no data. We thank Maria Lachgar-Ruiz for helpful comments on the manuscript text.

