## Additional File 1; Supplementary Figures 1-4 for "Investigating the Characteristics of Genes and Variants Associated with Self-Reported Hearing Difficulty in Older Adults in the UK Biobank"

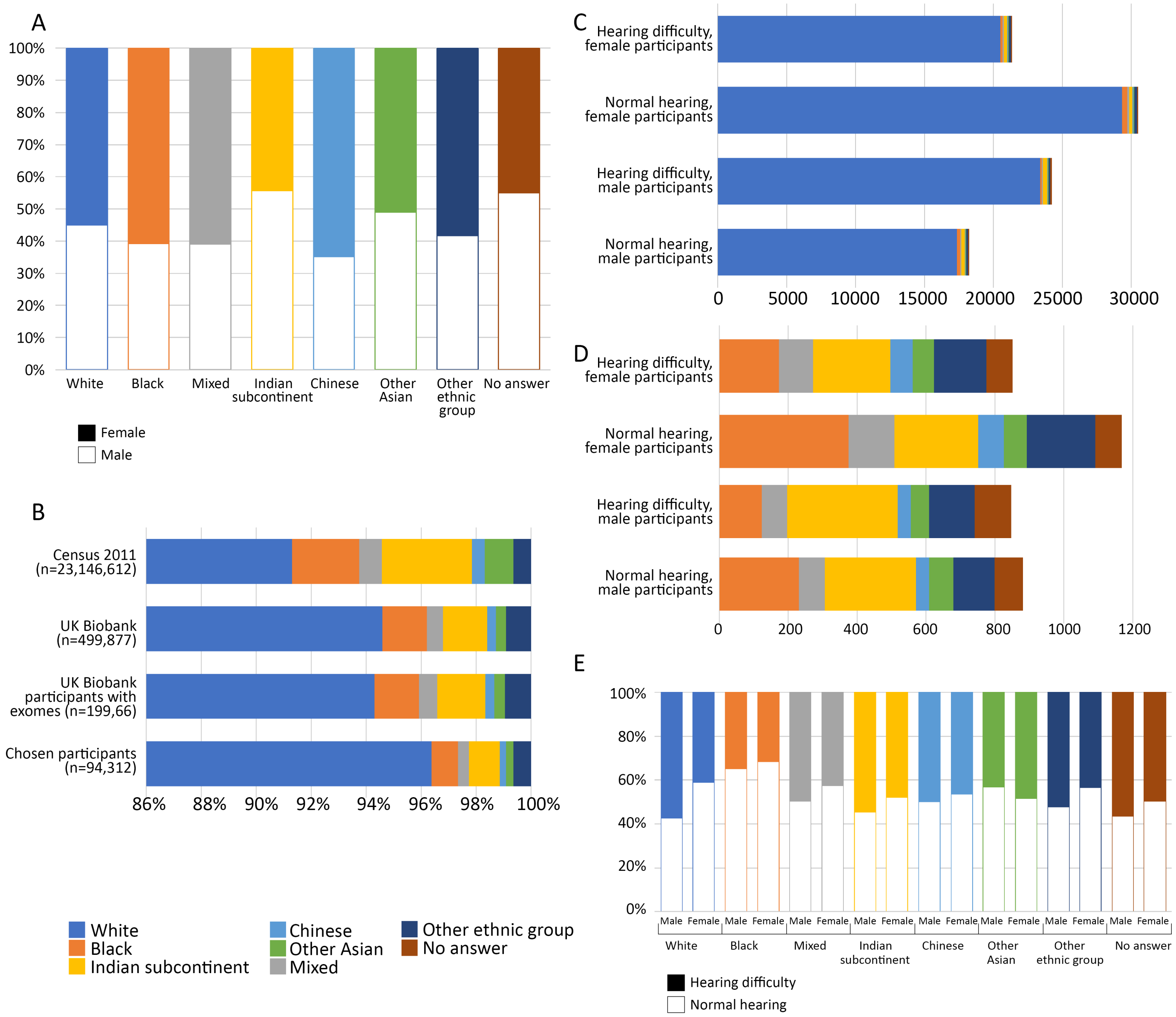


**Figure S1. Population characteristics of chosen participants.**

Bar charts showing numbers of people by sex, self-reported ethnicity and self-reported hearing phenotype. Ethnicities were combined to form broad groupings (see Additional File 2: Table S1). **A**, Percentage of male (empty bar) and female (filled bar) participants in each broad ethnic grouping in this study. **B**, Self-reported ethnicity in the UK census 2011 [56], the entire UK Biobank, UK Biobank participants with exomes, and this study, shown as a percentage of the total. Note that the x-axis starts at 86%, in order to show the minority percentages clearly. **C**, Self-reported ethnicity of people included in this study. **D**, Self-reported minority ethnicities of people included in this study. Note the difference in the range of the x-axis between C and D. **E**, Percentage of male and female participants classified in the self-reported hearing difficulty phenotype (filled bar) and classified in the self-reported normal hearing phenotype (empty bar) in each broad ethnic grouping.

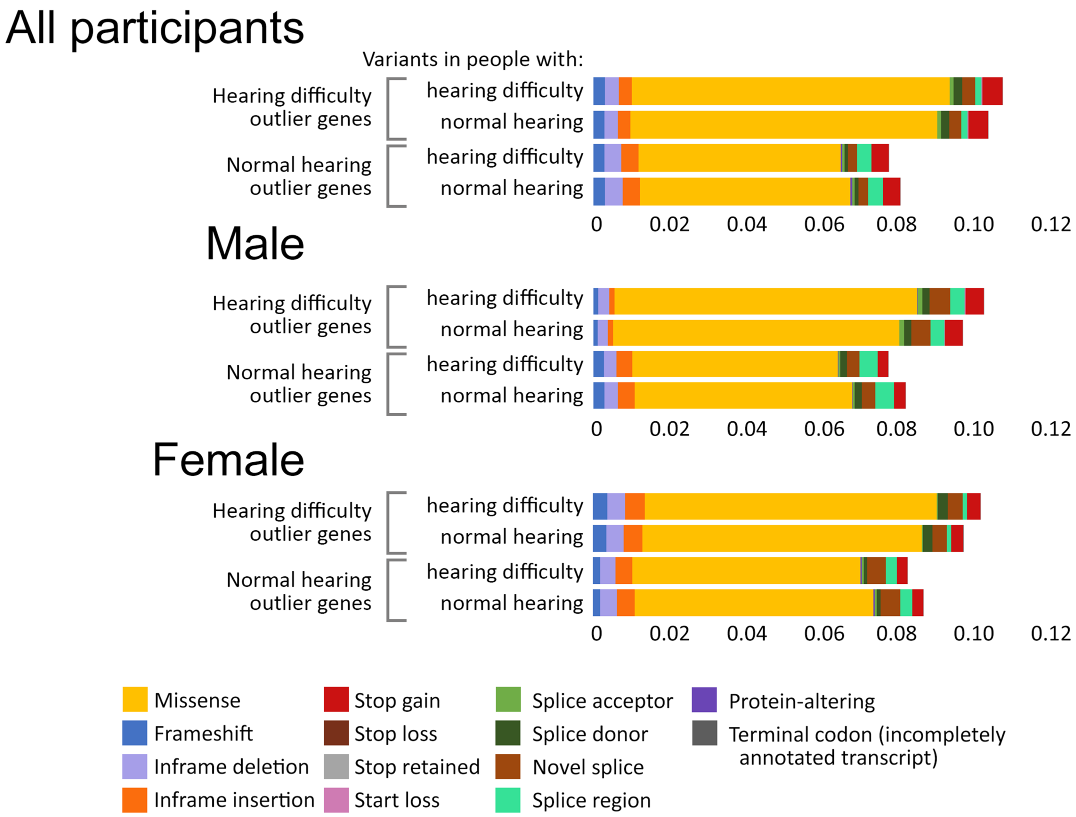


**Figure S2. Variant types in the outlier gene lists.**

Bar charts showing the number of variants per person, per gene, in the genes of the high variant load lists from high impact variants, MAF<0.1, coloured by maximum impact (see Additional File 1: Table S5). For each list of outlier genes, we have plotted the variants in people with hearing difficulty separately from those with normal hearing, to ask whether the variant impacts are different between the phenotype groups.

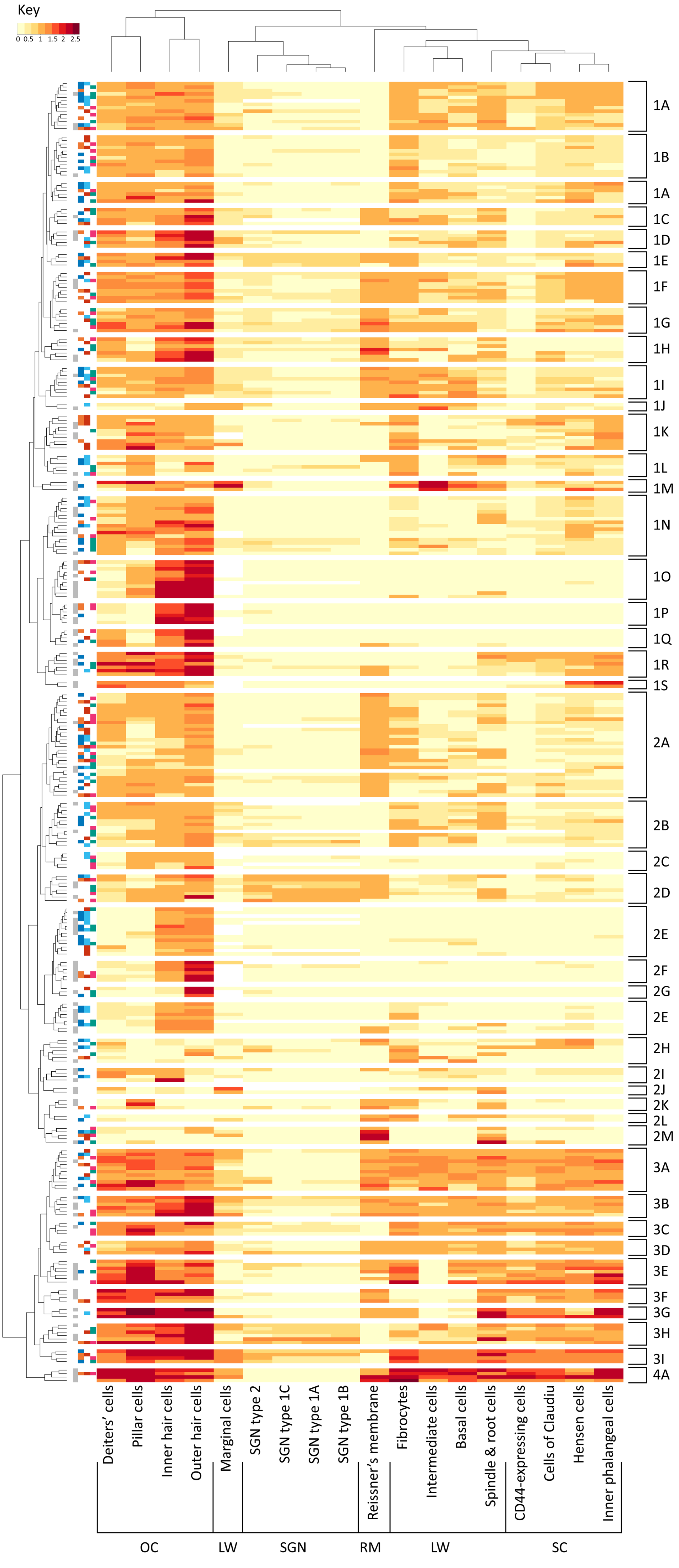
Key
0 0.5 1 1.5 2 2.5
1A
1B
1A
1C
1D
1E
1F
1G
1H
1I
1J
1K
1L
1M
1N
1O
1P
1Q
1R
1S
2A
2B
2C
2D
2E
2F
2G
2E
2H
2I
2J
2K
2L
2M
3A
3B
3C
3D
3E
3F
3G
3H
3I
4A
Deiters' cells
Pillar cells
Inner hair cells
Outer hair cells
Marginal cells
SGN type 2
SGN type 1C
SGN type 1A
SGN type 1B
Reissner's membrane
Fibrocytes
Intermediate cells
Basal cells
Spindle & root cells
CD44-expressing cells
Cells of Claudiu
Hensen cells
Inner phalangeal cells
OC
LW
SGN
RM
LW
SC

**Figure S3. Outlier and deafness gene heatmap.**

Heatmap showing maximum expression levels of the high quality one-to-one mouse orthologues of outlier and known deafness genes, across a range of inner ear cell types between E16 and P35. Cell types and genes are clustered according to their expression along the x and y axes respectively. We further defined gene clusters first based on the R dendrograms (taking the first three divisions, resulting in four main clusters numbered 1-4) and then based on the expression within specific cell types (subclusters labelled with letters). To annotate cell types, we grouped Deiters' cells, pillar cells, and inner and outer hair cells (as organ of Corti, OC), all spiral ganglion neuron types (as SGN), marginal, intermediate and basal cells, fibrocytes and spindle and root cells (as lateral wall, LW), and Hensen cells, cells of Claudius, inner phalangeal cells and CD44-expressing cells (CD44) as supporting cells (SC). Expression in Reissner's membrane was left as a single group (RM). The blocks of colour on the left denote deafness genes (grey, first column), outlier genes in normal hearing (blue) and hearing difficulty (orange) in all participants (second column), outlier genes in normal hearing (cyan) and hearing difficulty (red) in male participants (third column) and outlier genes in normal hearing (teal) and hearing difficulty (magenta) in female participants (fourth column). The genes are listed in the same order in Additional File 1: Table S8, along with their classifications and all annotations.

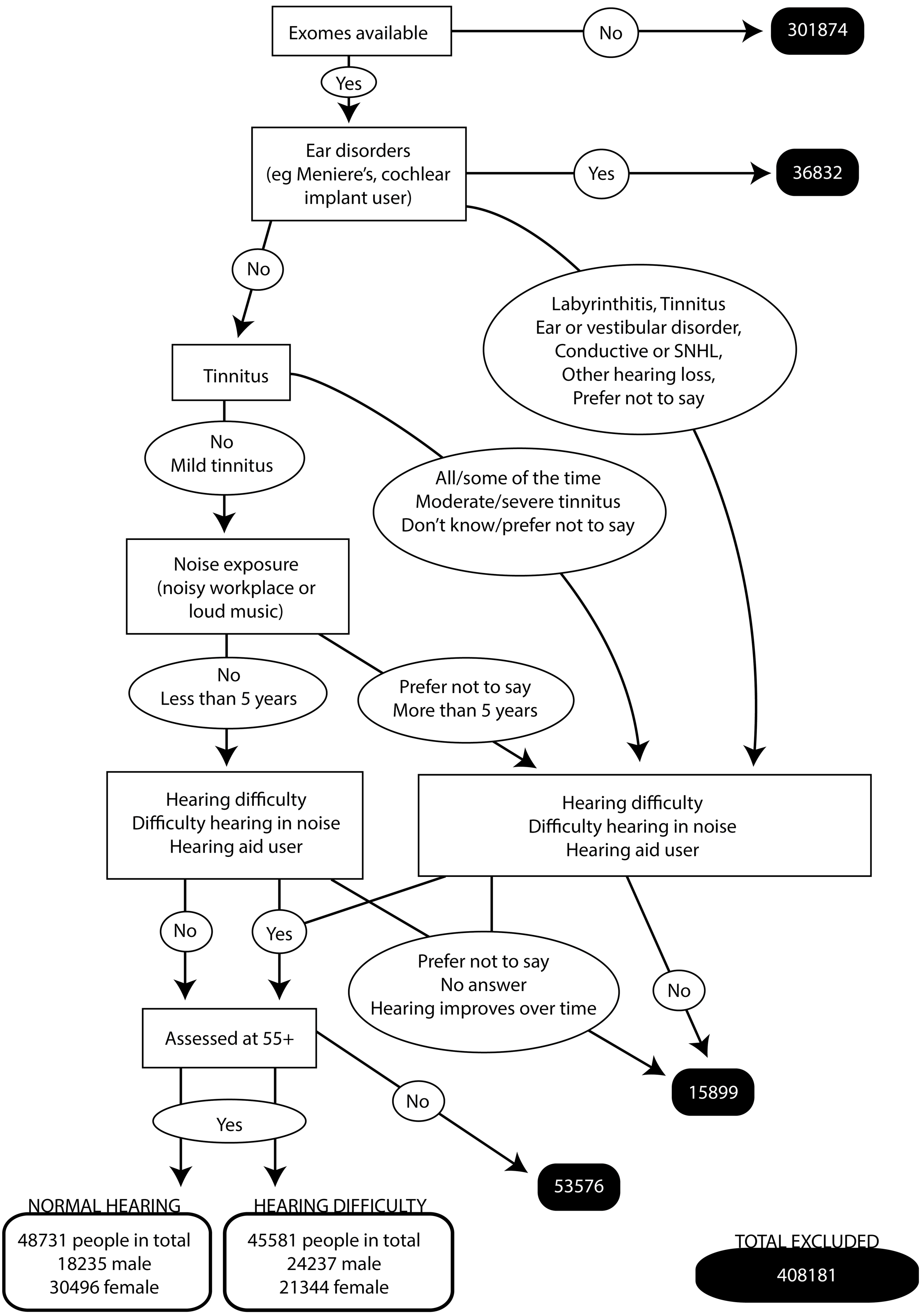


**Figure S4. Participant selection.**

Flow diagram showing the process of categorising participants by availability of exomes, their questionnaire answers, and age when assessed. In black boxes, the number of people excluded at each step.
